# Exposome-wide associations and polyexposure risks of workplace chemical exposures on adult asthma: The Personalized Environment and Genes Study (PEGS)

**DOI:** 10.1101/2025.05.02.25326863

**Authors:** Yujing Chen, Farida S. Akhtari, Yixuan He, Lihang Yu, Vy Kim Nguyen, Martin Tsz-Ki Tsui, Yuki Mizuno, William E. Funk, Crystal Ying Chan, Shelly LA Tse, Kin Fai Ho, Chirag J. Patel, Alison Motsinger-Reif, Ming Kei Chung

## Abstract

**Background:** Many potential occupational asthmagens and their co-exposure risks are underestimated. We conducted an exposome-wide association study (ExWAS) to systematically explore occupational chemical exposures and estimate their joint association with adult asthma and related multimorbidity.

**Methods:** This cross-sectional study included 3148 adults in the Personalized Environment and Genes Study (PEGS), a North Carolina-based cohort with questionnaire-based health and exposure data. Standardized questionnaires investigated occupational exposure to 93 chemical agents across 18 groups and the use of personal protective equipment (PPE). Current adult asthma was defined as self-reported doctor-diagnosed asthma with an attack in the prior year. Respiratory multimorbidity was ascertained if asthma coexists with allergic rhinitis or chronic obstructive pulmonary disease. Logistic regression-based ExWAS analyzed associations for individual chemicals. Weighted standardized polyexposure scores (PXS) were developed for predefined known asthmagens (PXS known) and suspected or novel risk agents (PXS suspected or novel) selected from ExWAS results through a regularized selection procedure.

**Results:** Approximately 9.7% of adults reported current asthma. In ExWAS, ever exposure to cleaning liquids, heavy metals, dust, and combustion emissions, was positively associated with asthma [adjusted odds ratio (aOR): 1.50-2.10; false discovery rate (FDR)< 0.05)], particularly when exposure occurred weekly or lasted over one year. The top five specific risk agents were talc, ammonia, bleach, silica, and carbon monoxide. PXS known (four agents) and PXS suspected or novel (six agents) showed positive associations with asthma and multimorbidity (aOR: 1.22-1.94 per 0.1 increment). Additionally, participants PPE had reduced odds of asthma and respiratory multimorbidity in relation to PXS suspected or novel (*P* interaction< 0.05).

**Conclusion:** We identified six suspected and novel chemical risk agents not yet recognized as occupational asthmagens by major classification systems. PXS can effectively summarize joint exposure effects on asthma and respiratory multimorbidity, supporting occupational risk assessment in clinical and public health settings.

## Introduction

Asthma is a chronic airway inflammation disease, affecting 262 million people worldwide in 2019.^1^ In adults, asthma commonly co-occurs with allergic rhinitis (AR) and chronic obstructive pulmonary disease (COPD), contributing to a great burden of symptoms, poor quality of life, and high mortality.^2–4^ Occupational exposure is an important factor in the environmental etiology of adult asthma.^5^ Approximately 16% of incident asthma is attributable to occupational exposures, and about 20% of workers with pre-existing asthma experience aggravated symptoms at work.^6,7^ However, the burden of work-related asthma may be underestimated, with a 31-year surveillance program reporting that only 48% of work-related asthma was due to recognized asthmagens. ^8^ This finding indicates that unknown chemical agents may account for a substantial portion of asthma cases.

To date, over 500 chemical agents have been identified as asthmagens by the World Allergy Organization (WAO)^9^ and the Association of Occupational and Environmental Clinics (AOEC).^10^ Asthmagen are further classified as either sensitizers or irritants. However, knowledge gaps remain in identifying new asthmagens and understanding their exposure patterns and co-exposure effects. ^11^ Recent studies in European populations have measured multiple occupational sensitizers and irritants using the occupational asthma-specific job exposure matrix (OAsJEM). ^12–14^ For example, a French cross-sectional study suggested that lifetime exposure to irritants including disinfectants/cleaning products and organic solvents was positively associated with current asthma in adults.^13^ Another French case-control study found that ever exposure to both occupational sensitizers and irritants was related to a higher risk of adult-onset asthma.^14^ However, the OAsJEM was developed based on hypothesized asthmagens in seven broad groups, and thus studies using this tool often focus on a limited set of exposure categories, restricting the ability to explore new asthmagens and characterize the risk associated with specific chemical agents.

Exposome-wide association studies (ExWAS) represent a hypothesis-free strategy that systematically assesses an array of exposures for their associations with a disease while minimizing selective reporting. ^15^ It is a standardized analytical approach to identify novel risk factors and serves as an initial screening for further in-depth analysis. ^16,17^ Particularly, polyexposure scores (PXS) were recently introduced to summarize the total effect of multiple independent exposures derived from ExWAS.^17^ While ExWAS-based analysis has been applied to investigate risk factors for asthma in certain domains such as urban environments^16^ and early-life factors^18^, it has not been employed to extensively explore occupational chemical asthmagens and their co-exposure effects.

Using a North Carolina, USA-based population, this study aimed to (1) systematically investigate the associations between occupational chemical exposures and adult asthma and respiratory multimorbidity using an ExWAS approach; and (2) develop weighted standardized PXS to assess the combined risk of known asthmagens and suspected or novel chemical agents for asthma outcomes. We also examined the effect modification by the use of personal protection equipment (PPE) to provide insights into potential intervention measures in workplaces and inform policy changes.

## Methods

### Study design and population

The Personalized Environment and Genes Study (PEGS) is a multi-ancestry cohort established in 2002 at the National Institute of Environmental Health Sciences (NIEHS). Participants (age> 18 years) were recruited from the general population at over 200 unique sites in North Carolina, such as healthcare clinics, hospitals, universities, and community centers. PEGS participants were administered three cross-sectional surveys to collect comprehensive information on health history, medication conditions, and environmental exposures. ^19^ From 2013 to 2022, 9,449 adults completed the Health and Exposure Survey, and 3,618 participated in the External Exposome Surveys (data version: Freeze 3.1). This study included 3,148 individuals with available data on asthma and occupational exposures. The PEGS study protocol was approved by the NIEHS institutional review boards, and all participants provided written informed consent. The Survey and Behavioural Research Ethics Committee at the Chinese University of Hong Kong approved this study protocol (SBRE-23-0352).

### Chemical exposures in the workplace

The list of investigated chemicals was determined based on the Agency for Toxic Substances and Disease Registry. ^20^ Occupational exposure was defined as contacting with unsealed chemical substances through inhalation, dermal contact, or ingestion in any job for at least 15 minutes a week. PEGS investigated over 100 chemical agents with survey questions asking participants whether they had ever been exposed to specific agents classified by chemical properties and/or industrial applications in 18 groups. We excluded unspecified agents within each group (i.e., “other”, n=18) and chemical agents with exposure in 20 or fewer individuals (n=22), resulting in 93 chemical agents (ever exposure, yes or no) in the final analysis. Exposure frequency (at least once per day/week/year) and duration (overall exposure years) to chemical groups were also collected.

We pre-classified chemicals as known asthmagens if they were identified by the AOEC or WAO. The AOEC Code list is the largest and most comprehensive catalog of occupational related asthmagens, with a formal review of chemical substances by occupational and pulmonary medicine experts since 2002.^10^ The WAO published a list of sensitizing agents for occupational asthma by summarizing previous findings up to 2019.^9^ **Table S1** displays chemical agents investigated in our study designated as known asthmagens. These agents were further categorized into sensitizers or irritants based on AOEC criteria and WAO-cited references. Chemical agents associated with asthma in this study but not in the AOEC or WAO list would be further considered suspected (supported by previous studies) or potential novel risk agents.

PEGS surveys also collected information on the use of PPE at work, including gloves, face mask or respirator, and protective clothing (e.g., Tyvek, lab coat, and apron). In addition, participants were asked about other chemical-related exposures, including renovation status and floor materials in the current or most recent workplaces.

### Asthma outcomes and respiratory comorbidity

Self-reported asthma diagnosis was determined by an affirmative answer to the question, “Has a doctor or other healthcare provider ever told you that you have asthma?” To confirm active asthma in adulthood, we further defined asthma cases if individuals 1) reported an asthma diagnosis, and 2) still had asthma or experienced asthma attacks, emergency room visits, or received medication prescriptions in the past 12 months, as outlined in **Figure 1 (A)**. Asthma control status in the past 14 days was measured by reported frequency of night waking, daily activity limitations, and symptoms like wheezing or tightness in the chest or cough due to asthma.

**Figure 1.**
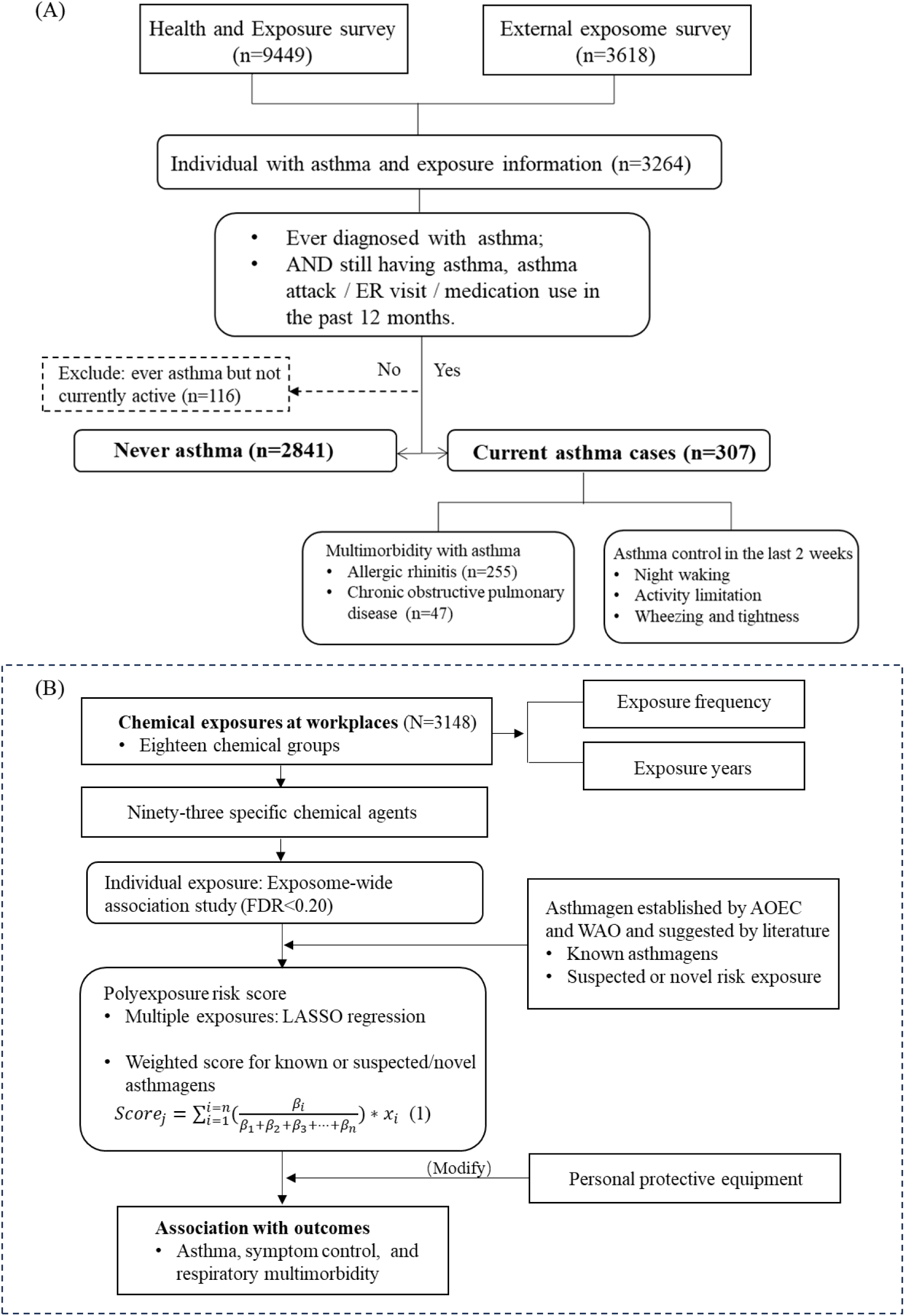
The inclusion of study population (A) and risk score design and analysis workflow (B) Abbreviations: AOEC, Association of Occupational and Environmental Clinics; WAO, World Allergy Organization. Weighted standardized scores on the basis of β coefficients of each chemical in the LASSO model were calculated with formula (1) Where subscript j refers to the values corresponding to jth participants; superscript i refers to the values corresponding to ith chemical, and *β* refers to the coefficient of each chemical in the logistic model; n refers to the total number of chemicals selected to generate the risk score.

Respiratory multimorbidity was defined as asthma and concurrent AR or COPD. AR was defined by a self-reported diagnosis of allergic rhinitis, hay fever, or seasonal allergies, and COPD was defined by self-reported diagnosis of chronic bronchitis or emphysema. PEGS survey questions were developed based on validated questionnaires in the Nurses’ Health Study II, National Health and Nutrition Examination Survey (NHANES), and the PhenX toolkit. ^21,22^

### Covariates

Due to the exploratory nature of the ExWAS approach, we included several key socio-demographic characteristics as covariates based on literature ^14,18^ and a directed acyclic graph (DAG) (**Figure S1**). The Health and Exposure survey collected participants’ age, sex, race, highest education level, annual household income, and BMI (kg/m^2^), which was calculated based on self-reported height and weight. Further details are provided in **Table 1**.

**Table 1.** General characteristics of study population in the PEGS (N=3148)

|  | Total<br>(N=3148) | Non-asthma<br>(n=2841) | Asthma<br>(n=307) | *p* value |
| --- | --- | --- | --- | --- |
| Age, year, Mean (SD) | 50.26 (14.39) | 50.38 (14.46) | 49.19 (13.66) | 0.171 |
| N-Miss | 20 | 19 | 1 |  |
| Sex, n (%) |  |  |  | **< 0.001** |
| Male | 945 (30.25) | **891 (31.58)** | **54 (17.82)** |  |
| Female | 2179 (69.75) | **1930 (68.42)** | **249 (82.18)** |  |
| N-Miss | 24 | 20 | 4 |  |
| Race, n (%) |  |  |  | **0.050** |
| White | 2622 (84.28) | **2378 (84.63)** | **244 (81.06)** |  |
| Black | 329 (10.58) | **285 (10.14)** | **44 (14.62)** |  |
| Others | 160 (5.14) | **147 (5.23)** | **13 (4.32)** |  |
| N-Miss | 37 | 31 | 6 |  |
| Highest education, n (%) |  |  |  | 0.558 |
| High school or below | 184 (5.86) | 164 (5.79) | 20 (6.51) |  |
| College | 359 (11.44) | 316 (11.16) | 43 (14.01) |  |
| Technical or vocational school graduate | 340 (10.83) | 305 (10.77) | 35 (11.40) |  |
| Bachelor | 1029 (32.78) | 934 (32.98) | 95 (30.94) |  |
| Graduate or professional | 1227 (39.09) | 1113 (39.30) | 114 (37.13) |  |
| N-Miss | 9 | 9 | 0 |  |
| Annual household income, n (%) |  |  |  | **< 0.001** |
| Less than \$20,000 | 186 (6.04) | **147 (5.30)** | **39 (12.75)** | |
| \$20,000 to 49,999 | 698 (22.65) | **626 (22.56)** | **72 (23.53)** | |
| \$50,000 to 79,999 | 853 (27.69) | **773 (27.86)** | **80 (26.14)** | |
| \$80,000 or above | 1344 (43.62) | **1229 (44.29)** | **115 (37.58)** | |
| N-Miss | 67 | 66 | 1 |  |
| BMI, kg/m<sup>2</sup>, Mean (SD) | 28.04 (6.71) | **27.75 (6.44)** | **30.70 (8.34)** | **< 0.001** |
| N-Miss | 25 | 24 | 1 |  |
| AR, n (%) |  |  |  | **< 0.001** |
| Yes | 1433 (45.52) | **1178 (41.46)** | **255 (83.06)** |  |
| No | 1985 (54.48) | **1663 (58.54)** | **52 (16.94)** |  |
| COPD, n (%) |  |  |  | **< 0.001** |
| Yes | 100 (3.18) | **53 (1.87)** | **47 (15.31)** |  |
| No | 3318 (96.82) | **2788 (98.13)** | **260 (84.69)** |  |

Abbreviations: AR, allergic rhinitis; COPD, chronic obstructive pulmonary disease; BMI, body mass index; PEGS, Personalized Environment and Genes Study.
Data were presented as mean ± SD or n (%).
P value from chis-square test or *t*-test.

### External dataset

We replicated our ExWAS findings in NHANES. A total of 14,174 adults who participated in the 2007-2008, 2009-2010, and 2011-2012 survey cycles were included. We selected comparable occupational chemical exposure groups in NHANES, including mineral dust, exhaust fumes, and other fumes. Further methodology details are provided in the **Supplementary Methods**.

### Statistical analysis

Characteristics of participants were described as means and standard deviations (SDs) for continuous variables or frequencies and percentages for categorized variables. Differences between asthma and non-asthma groups were compared using student *t*-tests or χ^2^ tests.

Two steps were taken to examine the individual and joint association between occupational chemicals and asthma-related outcomes, as shown in **Figure 1 (B)**. First, we fitted the logistic regression to assess the pairwise associations between each chemical group and specific agent with asthma and respiratory multimorbidity using the ExWAS approach. Exposure frequency (never, daily or weekly, monthly or yearly) and exposure duration (never, <1 year, 1∼<5 years, or ≥5 years) to chemical groups were examined in separate models. The Benjamini-Hochberg procedure was employed to control the false discovery rate (FDR) for multiple testing.

Second, weighted standardized PXS were generated based on the selected chemical agents. Using screening cut-off suggested by previous exposome studies ^16,18^, specific chemical agents with an FDR <0.20 in the ExWAS analysis were included in the multi-exposure analysis. The least absolute shrinkage and selection operator (LASSO, L1 regularization) penalized model was employed to select the most contributive variables. We used a 10-fold cross-validation approach, repeated 100 times, to determine the optimal degree of penalization. Specifically, known asthmagens were modeled as one set, while suspected and novel risk agents were modeled as the other set. Then, PXS _known_ (for known asthmagens) and PXS _suspected or novel_ (for chemical agents that are either suspected or novel risk factors) were calculated based on their coefficients (βs) from the LASSO regression using formula (1). A higher score indicates greater asthma risk from exposure to chemical agents (range: 0-1).

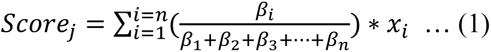

Where subscript *j* refers to participant *j*; superscript *i* refers to chemical *i*, and *β* refers to the coefficient of each chemical in the LASSO model; *n* refers to the total number of included chemicals.

We analyzed the associations of PXS with asthma and comorbid AR and COPD. The effect modification of PPE use on the observed significant association was examined by including the multiplicative interaction term. The main analytical model was adjusted for age, sex, race, highest education level, annual household income, and BMI based on the complete case data.

To test the reproducibility of ExWAS findings, associations between three chemical groups and asthma were examined in NHANES. Moreover, several sensitivity and supplementary analyses were performed in the PEGS cohort: 1) we imputed missing covariates data (missing rate: 0.03%-2.12%) with five iterations by employing Multivariate Imputation by Chained Equations; 2) we included other chemical agents with *P* < 0.05 or FDR < 0.30 in the ExWAS as covariates to examine whether the LASSO model consistently selected the chemical agents in the main analysis; 3) we considered PXS above the top 10% potentially high risk and reanalyzed the high-risk group in association with asthma and the effect modification of PPE; 4) we examined the association between PXS with asthma control measurements (night waking, daily activity limitations, and symptoms); 5) we analyzed whether sex modified the effect of chemical exposures; and 6) we explored association between workplace characteristics related to chemical exposures and asthma as supplementary results.

All analyses were performed in the R 4.2.0 (R Core Team, 2022) statistical software. The codes for this study can be found at https://github.com/Chenyj336/PEGS_chemical-exposome-and-asthma. Results can be interactively explored at https://lihangyu.shinyapps.io/asthma_shiny.

## Results

### General characteristics of the study population by asthma

**Table 1** shows that 3,148 participants in our study had a median age of 50.26 ± 14.39 years, with 69.75 % female and 84.28% White participants. A total of 307 (9.75%) adults reported current asthma, with a higher proportion being female and Black compared to those without asthma. Individuals with asthma also tended to have lower household income and higher BMI. Among asthma cases, 255 (83.06%) and 47 (15.31%) were concurrently diagnosed with AR and COPD, respectively.

### Workplace chemical exposures

In **Table 2**, for six chemical groups, exposure proportions exceeded 10%, with the highest frequencies observed for dyes and inks (26.68%), cleaning liquids (26.65%), and alcohols (20.17%). Among specific chemical agents (**Figure S2**), bleach was the most common exposure (24.68%). The exposure frequency and duration for 18 chemical groups are shown in **Figure S3,** and their pairwise correlations are depicted in **Figure S4.**

**Table 2.** Association of exposure to chemical groups in the workplace with asthma in PEGS participants (N=3148)

| Main chemical groups | n (%) | Crude model |  |  | Adjusted model |  |  |
| --- | --- | --- | --- | --- | --- | --- | --- |
|  |  | OR (95% CI) | *P* value | FDR | OR (95% CI) | *P* value | FDR |
| Solvents and degreasers | 469 (14.90) | 1.07 (0.77, 1.47) | 0.703 | 0.756 | 1.24 (0.88, 1.74) | 0.214 | 0.257 |
| Lubricating oils | 282 (8.96) | 1.21 (0.82, 1.78) | 0.345 | 0.517 | 1.44 (0.95, 2.20) | 0.088 | 0.144 |
| Cleaning liquids | 839 (26.65) | **1.67 (1.30, 2.14)** | **<0.001** | **0.001** | **1.50 (1.16, 1.95)** | **0.002** | **0.014** |
| Heavy metals | 180 (5.72) | 1.61 (1.05, 2.49) | 0.030 | 0.109 | **2.10 (1.33, 3.30)** | **0.001** | **0.013** |
| Alcohols | 635 (20.17) | 1.05 (0.78, 1.4) | 0.756 | 0.756 | 1.23 (0.91, 1.67) | 0.182 | 0.246 |
| Pesticides/Fumigants | 204 (6.48) | 1.59 (1.05, 2.4) | 0.028 | 0.109 | 1.60 (1.03, 2.48) | 0.038 | 0.109 |
| Compounds in plastic production | 129 (4.10) | 1.53 (0.92, 2.56) | 0.103 | 0.192 | 1.70 (1.00, 2.88) | 0.048 | 0.109 |
| Dust | 296 (9.40) | **1.72 (1.22, 2.43)** | **0.002** | **0.012** | **2.08 (1.45, 3.00)** | **<0.001** | **0.001** |
| Emission from fuels | 251 (7.97) | 1.45 (0.98, 2.13) | 0.060 | 0.180 | **1.73 (1.15, 2.60)** | **0.009** | **0.040** |
| Occupation Carcinogens | 421 (13.37) | 1.16 (0.83, 1.61) | 0.383 | 0.531 | 1.26 (0.89, 1.78) | 0.191 | 0.246 |
| Acids | 289 (9.18) | 0.87 (0.56, 1.33) | 0.508 | 0.653 | 0.98 (0.63, 1.53) | 0.931 | 0.931 |
| Alkalis | 210 (6.67) | 0.92 (0.56, 1.49) | 0.722 | 0.756 | 1.08 (0.65, 1.78) | 0.773 | 0.870 |
| Stains and varnishes | 198 (6.29) | 1.44 (0.93, 2.21) | 0.099 | 0.192 | 1.60 (1.00, 2.56) | 0.048 | 0.109 |
| Paints and paint thinners | 324 (10.29) | 1.31 (0.91, 1.87) | 0.144 | 0.236 | 1.43 (0.98, 2.08) | 0.066 | 0.118 |
| Anesthetics | 121 (3.84) | 1.54 (0.91, 2.61) | 0.107 | 0.192 | 1.78 (1.03, 3.07) | 0.038 | 0.109 |
| Glues and adhesives | 294 (9.34) | 1.38 (0.95, 1.99) | 0.087 | 0.192 | 1.32 (0.89, 1.94) | 0.169 | 0.246 |
| Soldering materials | 120 (3.81) | 0.84 (0.43, 1.61) | 0.594 | 0.712 | 1.06 (0.53, 2.12) | 0.870 | 0.921 |
| Dyes and inks | 840 (26.68) | **1.49 (1.16, 1.91)** | **0.002** | **0.012** | 1.29 (0.99, 1.69) | 0.057 | 0.115 |

Abbreviations: PEGS, Personalized Environment and Genes Study; OR, odds ratio; FDR, false discovery rate.
The crude logistic regression model was not adjusted for covariates.
The adjusted logistic regression model was adjusted for age, sex, race, highest education, annual household income, and BMI.

### ExWAS for chemical exposures and asthma outcomes

In the adjusted model (**Table 2)**, ever exposure to cleaning liquids (OR=1.50, 95%CI: 1.16, 1.95), heavy metals (OR=2.10, 95%CI: 1.33, 3.30), dust (OR=2.08, 95%CI: 1.45, 3.00), and emissions from the combustion of gasoline and other fuels (OR=1.73, 95%CI: 1.15, 2.60) were positively associated with asthma in adulthood at an FDR of 0.05 (*P*<0.009). **Figure 2** illustrates the ExWAS results for specific chemical agents, with an FDR of 0.20 (P<0.027) for 13 agents. The top five risk agents ranked by FDR were talc, ammonia, bleach, silica, and carbon monoxide.

**Figure 2.**
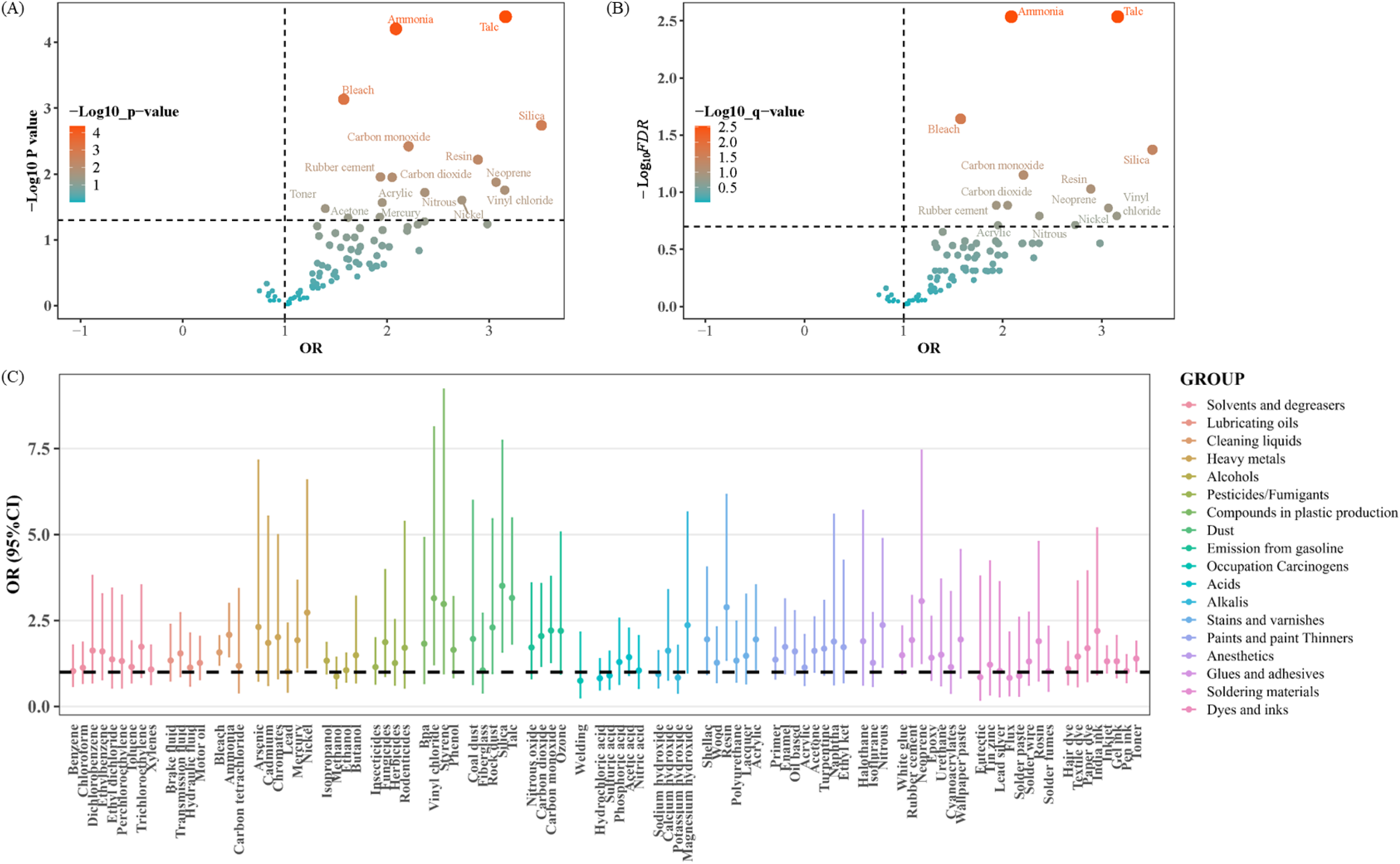
Association of specific chemical agents across 18 groups in the workplace with adult asthma (n=3148) (A) Volcano plot of the coefficient estimates and *P* value in the ExWAS analysis. The dashed horizontal line represents *P* =0.05; the dashed vertical line represents *OR*=1.00. The exposures with the *P* value<0.05 are labeled. (B) Volcano plot of the coefficient estimates and *false discovery rate (FDR)* in the ExWAS analysis. The dashed horizontal line represents *FDR* =0.20; the dashed vertical line represents *OR*=1.00. FDR: *P* values adjusted for the multiple tests according to the Benjamini & Hochberg procedure, the expected proportion of false discoveries amongst the rejected hypotheses. The exposures with FDR<0.20 are labeled. (C) Forest plot for the association of individual chemical exposure and asthma. The dotted horizontal line represents *OR*=1. Abbreviations: ExWAS= exposome-wide association study; OR =odds ratio. The ExWAS analysis was adjusted for age, sex, race, highest education, annual household income, and BMI.

Chemical groups were more likely to show significant association with asthma when exposures occurred daily or weekly or lasted over one year (**Table S2**). The estimated ORs for all specific chemical agents included in the ExWAS are presented in **Table S3.** We found overlapping risk chemical exposures for asthma and respiratory multimorbidity (**Table S4-S6**).

### PXS for multiple chemical agents and the effect modification of PPE

**Table 3** presents LASSO coefficients used to generate PXS based on four known asthmagens and six suspected or potential novel risk agents (FDR<0.20). As illustrated in **Figure 3** and **Table S7**, a 0.1 increase in PXS _known_ and PXS _suspected or novel_ was associated with an OR of 1.19 (95% CI: 1.11, 1.28) and 1.44 (95% CI: 1.27, 1.65) for asthma. Adults who used PPE had lower odds of asthma and comorbid AR and COPD when exposed to PXS _suspected or novel_. For example, a 0.1 increase in PXS _suspected or novel_ was associated with a 1.27-fold (95% CI: 1.27, 1.49) higher odds of asthma for adults who wore gloves, compared with a 2.01-fold (95% CI: 1.57, 2.57) higher odds for those who did not (*P* _interaction_<0.05). Similar moderating effects of protective clothing were observed for asthma and comorbid AR. No modifying effects of protective measures were found for the PXS _known_-related risk of asthma outcomes.

**Figure 3.**
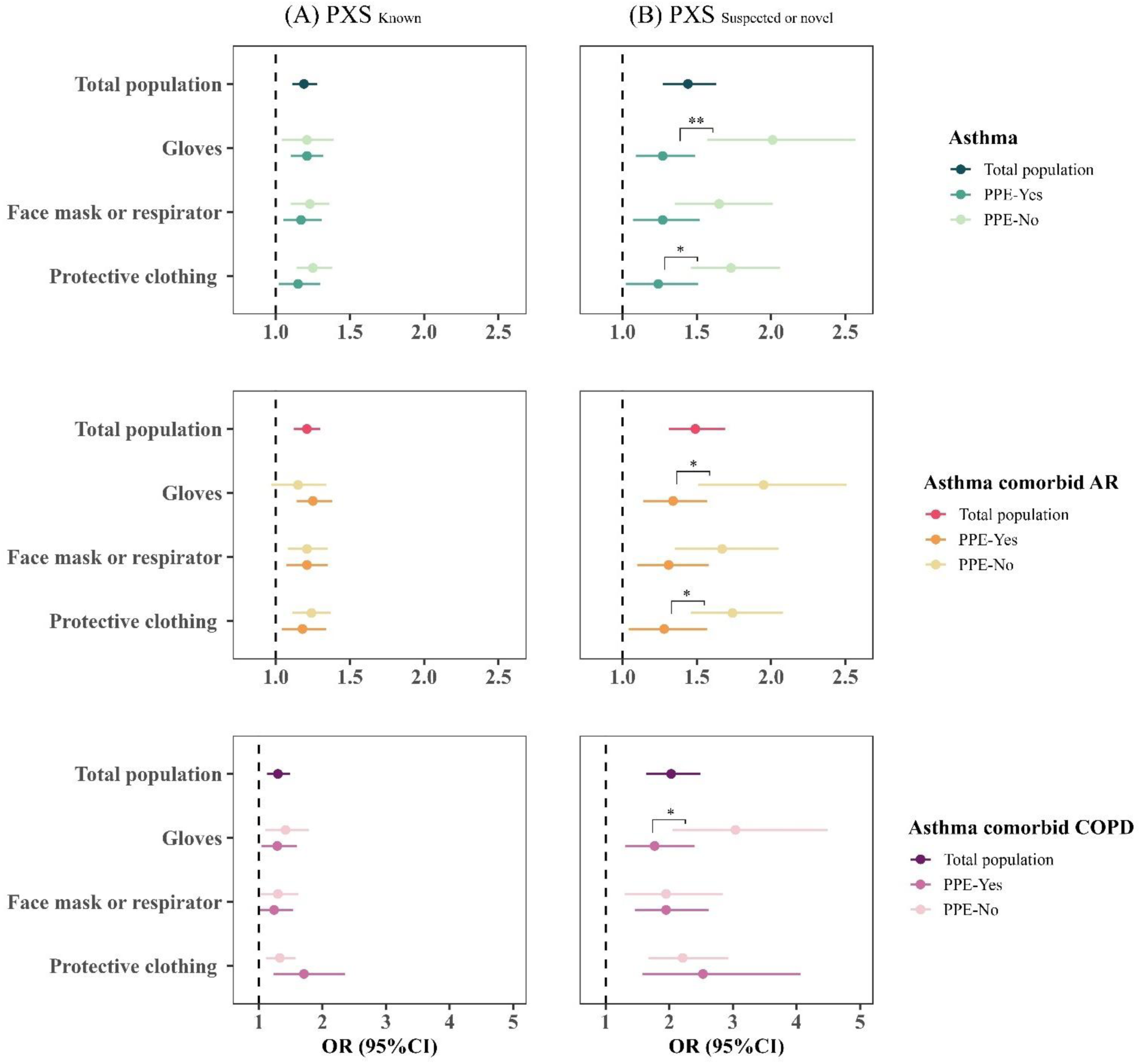
Estimated odds ratios (ORs) for asthma and respiratory multimorbidity in association with per 0.1 increase in polyexposure risk scores (PXS) of (A) known asthmagens and (B) suspected or novel risk agents. Effect modifications of protective measures including gloves, face mask, and protective clothing in adults are also shown. Abbreviations: AR, allergic rhinitis; COPD, chronic obstructive pulmonary disease; PPE, personal protective equipment. * *P* _interaction_< 0.05; ** *P* _interaction_ < 0.01 The logistic model was adjusted for age, sex, race, highest education level, annual household income, and BMI. Weighted standardized PXS (range: 0 to 1) were calculated based on the *β* coefficients of chemical exposures in the LASSO regression.

**Table 3.**
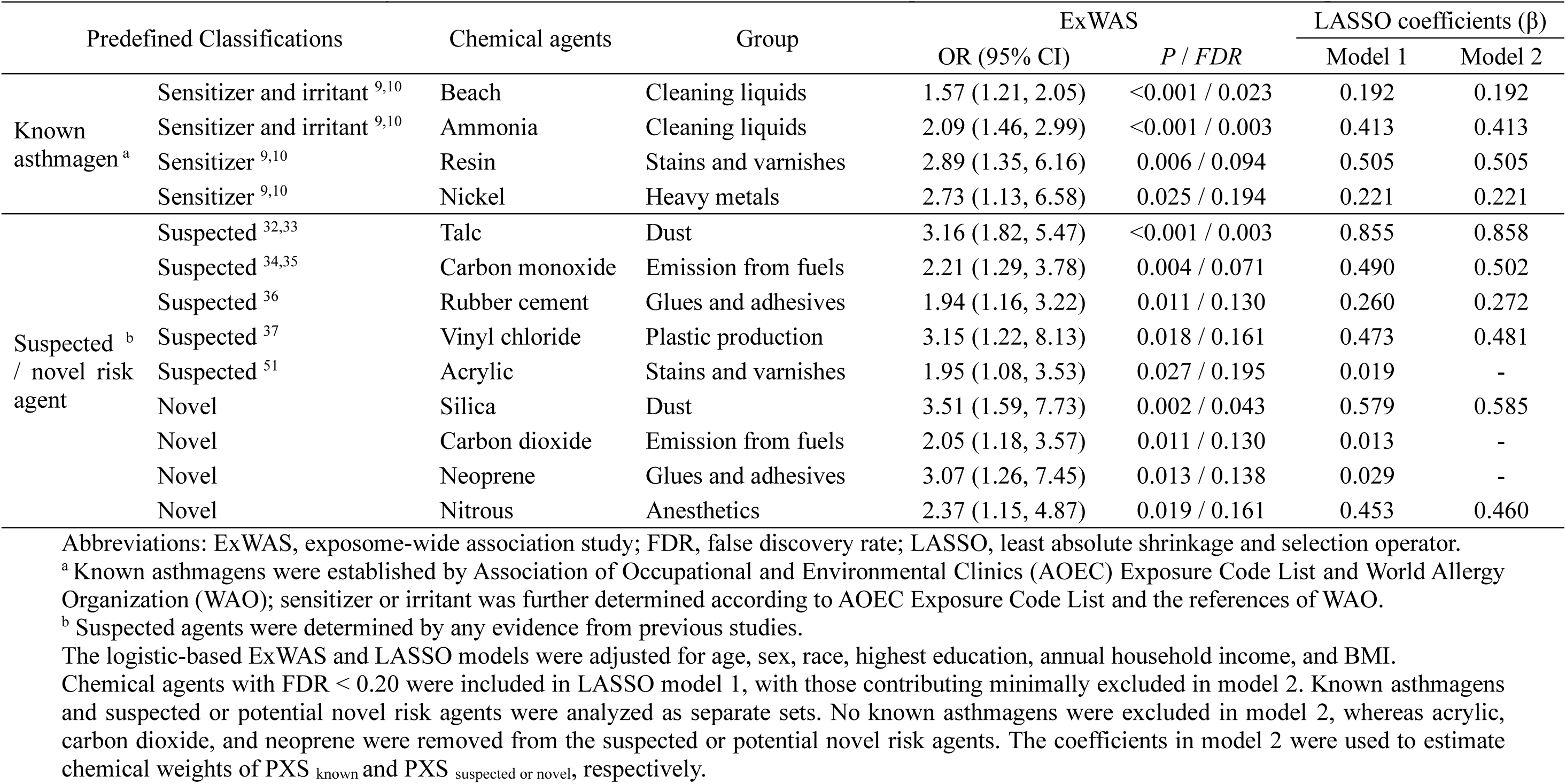
Potential risk chemical agents selected from ExWAS results and their multi-exposure model for adult asthma in the workplace (n=3148)

| Predefined Classifications | | Chemical agents | Group | ExWAS | | LASSO coefficients ($\beta$) | |
| --- | --- | --- | --- | --- | --- | --- | --- |
|  |  |  |  | OR (95% CI) | *P* / *FDR* | Model 1 | Model 2 |
| Known<br>asthmagen<sup>a</sup> | Sensitizer and irritant<sup>9,10</sup> | Beach | Cleaning liquids | 1.57 (1.21, 2.05) | <0.001 / 0.023 | 0.192 | 0.192 |
|  | Sensitizer and irritant<sup>9,10</sup> | Ammonia | Cleaning liquids | 2.09 (1.46, 2.99) | <0.001 / 0.003 | 0.413 | 0.413 |
|  | Sensitizer<sup>9,10</sup> | Resin | Stains and varnishes | 2.89 (1.35, 6.16) | 0.006 / 0.094 | 0.505 | 0.505 |
|  | Sensitizer<sup>9,10</sup> | Nickel | Heavy metals | 2.73 (1.13, 6.58) | 0.025 / 0.194 | 0.221 | 0.221 |
| Suspected<sup>b</sup><br>/ novel risk<br>agent | Suspected<sup>32,33</sup> | Talc | Dust | 3.16 (1.82, 5.47) | <0.001 / 0.003 | 0.855 | 0.858 |
|  | Suspected<sup>34,35</sup> | Carbon monoxide | Emission from fuels | 2.21 (1.29, 3.78) | 0.004 / 0.071 | 0.490 | 0.502 |
|  | Suspected<sup>36</sup> | Rubber cement | Glues and adhesives | 1.94 (1.16, 3.22) | 0.011 / 0.130 | 0.260 | 0.272 |
|  | Suspected<sup>37</sup> | Vinyl chloride | Plastic production | 3.15 (1.22, 8.13) | 0.018 / 0.161 | 0.473 | 0.481 |
|  | Suspected<sup>51</sup> | Acrylic | Stains and varnishes | 1.95 (1.08, 3.53) | 0.027 / 0.195 | 0.019 | - |
|  | Novel | Silica | Dust | 3.51 (1.59, 7.73) | 0.002 / 0.043 | 0.579 | 0.585 |
|  | Novel | Carbon dioxide | Emission from fuels | 2.05 (1.18, 3.57) | 0.011 / 0.130 | 0.013 | - |
|  | Novel | Neoprene | Glues and adhesives | 3.07 (1.26, 7.45) | 0.013 / 0.138 | 0.029 | - |
|  | Novel | Nitrous | Anesthetics | 2.37 (1.15, 4.87) | 0.019 / 0.161 | 0.453 | 0.460 |

Abbreviations: ExWAS, exposome-wide association study; FDR, false discovery rate; LASSO, least absolute shrinkage and selection operator.
<sup>a</sup> Known asthmagens were established by Association of Occupational and Environmental Clinics (AOEC) Exposure Code List and World Allergy Organization (WAO); sensitizer or irritant was further determined according to AOEC Exposure Code List and the references of WAO.
<sup>b</sup> Suspected agents were determined by any evidence from previous studies.
The logistic-based ExWAS and LASSO models were adjusted for age, sex, race, highest education, annual household income, and BMI.
Chemical agents with FDR < 0.20 were included in LASSO model 1, with those contributing minimally excluded in model 2. Known asthmagens and suspected or potential novel risk agents were analyzed as separate sets. No known asthmagens were excluded in model 2, whereas acrylic, carbon dioxide, and neoprene were removed from the suspected or potential novel risk agents. The coefficients in model 2 were used to estimate chemical weights of PXS<sub>known</sub> and PXS<sub>suspected or novel</sub>, respectively.

### Sensitivity and supplementary analysis

In **Table S8** and **Table S9,** similar results were observed when using imputed covariates or adjusted for other chemical agents. We consistently found that participants with the top 10 % of PXS _known_ and PXS _suspected or novel_ had higher odds of asthma and multimorbidity in **Table S10**. Both PXS _known_ and PXS _suspected or novel_ were associated with poor asthma control measurements (**Table S11**). The positive association between PXS and asthma was more robust and pronounced in females than in males (**Table S12**). However, we did not observe an association between recent renovations and floor material in the workplace a (**Table S13**).

### Replication in the NHANES

**Table S14** presents the characteristics of NHANES study populations. In **Table S15**, ever exposure to mineral dust (OR=1.23, 95%CI: 1.01, 1.50) and other fumes (OR=1.25, 95%CI 1.05, 1.48) were associated with increased odds of asthma in the combined survey cycles. Exposure to exhaust fumes showed a positive trend in association with asthma (*P*=0.076).

## Discussion

This study showed that adulthood exposure to various occupational chemical hazards, particularly cleaning liquids, heavy metals, dust, and combustion emissions, was associated with higher odds of asthma. Specifically, four known asthmagens and six chemical agents that are either suspected or novel risk factors were identified in this study. PXS for these substances demonstrated positive associations with asthma and comorbid AR and COPD. Reduced risks of asthma and respiratory multimorbidity was observed for individuals who used PPE when exposed to suspected or novel risk agents, providing implications for preventive practices in occupational settings.

Our findings reinforce previous understanding of certain asthmagens listed by the AOEC and WAO. Cleaning agents, which were among the most common exposures in our study (26.65%), are also reported as the most prevalent causes of work-related asthma across all industries in the U.S.^8^ This group of chemicals can cause excessive exposure to harmful volatile substances and act as irritants or sensitizers. ^23^ We identified bleach and ammonia as predominant hazardous cleaning agents linked to asthma risk, consistent with previous findings.^24^ Bleach, a chlorine-releasing agent, can liberate chloramines when mixed with ammonia solutions. Chlorine has been documented to have irritant properties that provoke airway hyperresponsiveness, oxidative stress, and neutrophilic inflammation, even at low doses in animal studies.^25,26^ An epidemiological study using the OAsJEM also indicated a deleterious effect of low to moderate chronic exposure to disinfectants and cleaning products on asthma risk.^13^ We found that more frequent or prolonged exposure in adulthood had a more pronounced association with asthma. These findings underscore the importance of reducing ubiquitous workplace exposure to cleaning agents to minimize asthma risk.

This study also identified two specific low molecular weight (LMW, <10 kDa) sensitizers in the groups of 1) heavy metals and 2) stains and vanishes, which are often reactive chemicals compared to high molecular weight allergens. ^27^ Contact allergy to nickel is prevalent among the general population ^28^, which was also associated with respiratory allergies including asthma.^29^ In occupational settings, a dose-response relationship between nickel exposure levels and a decline in lung function has been consistently reported. ^30^ Similar associations with occupational allergies were also observed for resin. The epoxy resin system, where resin is mixed with a hardener (e.g., phthalic anhydride derivatives and triglycidyl isocyanurate), was suggested to be highly reactive and cause IgE-mediated skin sensitization, with recent studies increasingly recognizing its respiratory sensitization potential. ^31^

Several potential suspected or novel risk factors not listed as asthmagens, mainly from the groups of dust, combustion emissions from fuels, and plastic production, were determined using the data-driven approach in this study. Among these, talc dust was the top risk agent in the ExWAS analysis and was linked to asthma-related outcomes in two case reports. One study found that inhalation of household cosmetic talc was related to intense endobronchitis and new-onset wheezing^32^, while another suggested that talc was responsible for poorly controlled asthma and unexplained bronchiolitis in a dental technician.^33^ Additionally, carbon monoxide, vinyl chloride, and rubber cement were also linked to asthma risk in this study. Specifically, ambient carbon monoxide was associated with declined lung function in asthmatic adults^34^ and increased asthma hospitalization risk^35^, but related evidence in occupational settings is lacking. The observed association for vinyl chloride may be attributable to its co-exposure with polyvinyl chloride, a known asthma sensitizer released during the plastic production process.^36^ For rubber cement, its inhaled chemical compositions (e.g., toluene) have been investigated in association with asthma symptoms and immune function with inconsistent findings.^37^ In addition to these suspected agents, our study also provides clues for two novel risk chemicals of asthma, including silica and nitrous, whose relationships with asthma remain ambiguous. The findings of independent NHANES dataset consistently support mineral dust (e.g., sand, coal, asbestos, silica) as a potential asthma risk factor, with fumes from fuel combustion and other sources also showing suggestive positive associations. These findings warrant confirmation in future occupational cohorts.

To evaluate the combined effect of multiple exposures, we generated PXS based on a regularized selection procedure while considering inter-exposure correlation. Limited studies have examined the joint effect of multiple chemical agents on asthma, with some using the clustering analysis to identify co-exposure patterns ^18,38^ and others employing the OAsJEM to explore the classifications of sensitizers and irritants.^14^ PXS are as a powerful tool to predict disease risk by aggregating the effects of multiple environmental exposures.^17,39^ We found significant positive associations between calculated standardized PXS and asthma and its control status. Consistent associations were observed for respiratory multimorbidity, which suggests that asthma and these concurrent conditions may share common risk agents and etiologies. Notably, the main contributors with large weights in the PXS (i.e., resins, cleaning agents, talc, and silica) have been shown to increase the risk of AR-related symptoms, chronic bronchitis, and declined lung function. ^31,40–42^ AR often precedes asthma due to the link between upper and lower airway allergies and inflammation, while asthma-COPD overlap is common in older adults with persistent airway limitation. ^4^ Given that individuals with asthma and comorbidities tend to experience frequent asthma exacerbations and a heavy disease burden ^2–4^, preventive measures should prioritize controlling shared risk exposures for respiratory multimorbidity in the workplace.

The effectiveness of PPE in preventing occupational respiratory diseases is inconsistent, possibly because PPE use often coincides with high exposure levels or the presence of other control measures. ^43^ In our study, we found that the use of gloves, protective clothing, and a face mask or respirator helped alleviate the risk of asthma and multimorbidity in relation to PXS _suspected or novel_. PPE may reduce exposure levels and contact allergies, but its effectiveness may depend on the characteristics of hazards and their exposure pathways. Some previous studies have suggested that PPE may not offer sufficient protection for asthma risk in miners and construction workers.^44,45^ However, our results provide evidence regarding multiple occupational exposures (e.g., dust, combustion emissions, and plastic production), supporting studies that found the beneficial effect of PPE.^46,47^ Nevertheless, PPE can serve as a last line of defense against occupational hazards when engineering or administrative controls are not feasible or effective.^48^ Given the low PPE usage rate (24.1-52.1% in our study), its use should be further promoted and incorporated into comprehensive workplace intervention programs.

This study has the strength of systematic investigation using a large set of occupational chemical agents, enabling novel findings with reduced reporting bias. However, we acknowledge several limitations. First, asthma diagnosis and chemical exposures were self-reported via standardized questionnaires, which may lead to underestimated prevalence and biased effect estimates toward the null. Nonetheless, data from semi-quantitative questions allowed us to assess chronic exposure to many chemical agents in any job held in adulthood, and our results were comparable to previous questionnaire-based studies.^38,49^ Second, this study may lack sufficient statistical power to detect risk chemicals with very low exposure frequency in the general population. For example, while we found suggestive evidence for some chemical groups (e.g., pesticides or fumigants and paints and paint thinners), we did not identify specific risk agents within these classes. Third, due to the exploratory nature of ExWAS and the cross-sectional study design, we could not draw causal conclusions based on the observed associations. However, we replicated our findings on mineral dust and fumes in NHANES, although comprehensive workplace exposure data were not available in current publicly accessible datasets. Additional prospective cohorts are needed to validate our findings while controlling for additional confounders. Fourth, our study population had a higher proportion of women (69.75%), and caution should be exercised when generalizing our findings. Females were more likely to have asthma and more vulnerable to suspected or novel chemical agents in our sensitivity analysis, which is consistent with literature suggesting women are more susceptible to adult-onset asthma.^50^ This may also help explain the smaller effect estimates we observed in the replication dataset, which had a more balanced sex ratio (50.52% female).

To conclude, this study used a systematic strategy to discover novel risk chemical agents for adult asthma, highlighting the potential adverse impacts of cleaning liquids, heavy metals, dust, and combustion emissions in the workplace. The estimated PXS showed a detrimental association with asthma and concurrent AR and COPD. Occupational clinicians and health care professionals should consider these chemical agents and promote the use of PPE in workers to mitigate the risk of asthma-related multimorbidity.

## Data Availability

All data produced in the present study are available upon reasonable request to the authors

## Acknowledgements

This study was supported by a grant from the Chinese University of Hong Kong (4937217).

## Author contributions

Y.C. and M.K.C. contributed to study conception and results interpretations. Y.C. and L.H. contributed to data cleaning, formal analysis, and visualization. F.S.A. and A.M. contributed to PEGS design, data collection and curation, and quality assurance. Y.C. drafted the manuscript. V.K.N., M.T.T., Y.M., W.E.F., C.Y.C., S.L.T., K.F.H., and C.J.P., contributed to methodology and critical revisions of the manuscript. M.K.C. was responsible for project supervision and funding. All authors reviewed the manuscript, gave insightful comments, and gave their approval.

## Data availability

The authors declare that the data supporting the findings of this study are available within the paper and its Supplementary Materials. The data will be available under controlled access to comply with polices and process of the PEGS data and meet applicable NIH requirements for data sharing and that the privacy and confidentiality of human subjects are protected. Researchers seeking access to PEGS data may contact the [PEGS Executive Leadership Committee](https://www.niehs.nih.gov/research/atniehs/labs/crb/studies/pegs/collaboration/proposal) with details from the PEGS websites.

