## Supplementary Methods for "Exposome-wide associations and polyexposure risks of workplace chemical exposures on adult asthma: The Personalized Environment and Genes Study (PEGS)"

**Contents**

**Supplementary Methods**

**Supplementary Tables and Figures**

**Table S1** Occupational asthmagens identified by AOEC and WAO in PEGS study (n=3148)

**Table S2** Association of exposure frequency and duration of main chemical exposure groups in the workplace with asthma in adults (N=3148)

**Table S3** Association of exposure to specific chemical agents in the workplace with asthma in adults (n=3148)

**Table S4** Association between exposure to main chemical groups and multimorbidity in adults (n=3148)

**Table S5** Association of exposure to specific chemical agents in the workplace with multimorbidity (asthma and AR) in adults (n=3148)

**Table S6** Association of exposure to individual chemicals in the workplace with multimorbidity (asthma and COPD) in adults (n=3148)

**Table S7** Estimated odd ratios (ORs) for asthma and respiratory multimorbidity in association with polyexposure risk scores (PXS) of known asthmagens and suspected or novel risk agents and the effect modification of PPE in adults (**data for Figure 3**)

**Table S8** ExWAS results for potential risk chemical agents and their multi-exposure model for adult asthma in the workplace with imputed covariates data (n=3148)

**Table S9** Additionally adjusted multi-exposure models for chemical agents and adult asthma in the workplace (n=3148)

**Table S10** Estimated odd ratios (ORs) for asthma and respiratory multimorbidity in association with high risk (top 10% PXS) of known asthmagens and suspected suspected/novel risk agents and the effect modification of PPE in adults

**Table S11** Association of polyexposure risk scores (PXS) of chemical exposures and related protective measures with asthma symptom control and respiratory multimorbidity in patients with asthma (N=307)

**Table S12** The modification effect of sex on the association of polyexposure risk scores (PXS) with asthma and respiratory multimorbidity in adults

**Table S13** Workplace characteristics and asthma and respiratory multimorbidity in adults

**Table S14** Demographic characteristics, asthma, and work environment exposures and in the U.S. adults, NHANES 2007–2012 (n = 14174)

**Table S15** Association between asthma and work environment exposures and in the U.S. adults, NHANES 2007–2012 (n = 14174)

**Figure S1** Directed acyclic graph (DAG) for the hypothesized relationship between occupational chemical exposures and adult asthma in the PEGS participants

**Figure S2** Proportion of adults ever exposure to individual chemicals within eighteen groups in the workplace (n=3148)

**Figure S3** Exposure frequency (A) and duration (B) to chemical groups at work in adults (n=3148)

**Figure S4** Pairwise correlation among chemical exposure groups

### Supplementary Methods

#### Study population

The NHANES is a nationally representative survey of the civilian, non-institutionalized U.S. population conducted by the Centers for Disease Control and Prevention (CDC). We used data from the 2007-2008, 2009-2010, and 2011-2012 NHANES cycles. Inclusion criteria are (1) participants aged 18 years or above; (2) complete asthma survey data; and (3) available occupational exposure data comparable to those investigated in the PEGS cohort. In total, 14,174 adults with complete asthma, occupational chemical exposure, and covariate data were included in the final analysis. The study protocol was approved by the National Center for Health Statistics Research Ethics Review Board.

#### Asthma and workplace exposure measurements

Current asthma in adults was investigated using similar questions in the NHANES. We defined asthma cases as individuals who 1) reported an asthma diagnosis, and 2) still had asthma or experienced either an asthma attack, emergency room visits, or doctor-prescribed medication in the past 12 months.

Although specific chemical agent exposures were not directly measured, three occupational exposure categories were investigated, which were comparable to those investigated in the PEGS cohort. Participants were asked the questions: "in any job, have you ever been exposed to mineral dust/exhausted fume/other fumes?" Mineral dust was defined as dust from rock, sand, concrete, coal, asbestos, silica, or soil. Exhaust fumes were described as exhaust fumes from trucks, buses, heavy machinery, or diesel engines. Other fumes refer to any other gases, vapors or fumes, such as vapors from paints, cleaning products, glues, solvents, and acids; or welding/soldering fumes.

#### Statistical analysis

Survey-weighted generalized linear regression with a logit link was employed to assess the association between asthma and workplace chemical exposures. The model was adjusted for age, sex, race/ethnicity, the highest education level, annual household income, BMI, and survey cycle, as detailed in **Table S14**.

### Supplementary Tables and Figures

**Table S1** Occupational asthmagens identified by AOEC and WAO in PEGS study (n=3148)

See Supporting Table S1 that in the .xlsx format

**Table S2** Association of exposure frequency and duration of main chemical exposure groups in the workplace with asthma in adults (N=3148)

| | Exposure frequency [a] | | | | | | Exposure years [a] | | | | | | | | |
|---|---|---|---|---|---|---|---|---|---|---|---|---|---|---|---|
| | Daily or weekly | | | Monthly or yearly | | | <1 year | | | 1~<5 years | | | >=5 years | | |
| **Main groups** | OR (95%CI) | *P* | FDR | OR (95%CI) | *P* | FDR | OR (95%CI) | *P* | FDR | OR (95%CI) | *P* | FDR | OR (95%CI) | *P* | FDR |
| Solvents and degreasers | 1.34 (0.89, 2.01) | 0.159 | 0.381 | 1.24 (0.68, 2.26) | 0.492 | 0.681 | 1.55 (0.72, 3.36) | 0.267 | 0.577 | 1.21 (0.8, 1.83) | 0.365 | 0.731 | 0.99 (0.44, 2.22) | 0.987 | 0.987 |
| Lubricating oils | 1.50 (0.83, 2.70) | 0.183 | 0.412 | 1.67 (0.86, 3.26) | 0.130 | 0.350 | 1.66 (0.91, 3.06) | 0.100 | 0.372 | 1.18 (0.6, 2.32) | 0.626 | 0.811 | 1.29 (0.36, 4.6) | 0.698 | 0.811 |
| Cleaning liquids | **1.73 (1.29, 2.31)** | **<0.001** | **0.004** | 1.27 (0.73, 2.22) | 0.390 | 0.639 | 1.32 (0.81, 2.14) | 0.258 | 0.577 | **1.62 (1.18, 2.23)** | **0.003** | **0.058** | 1.56 (0.87, 2.78) | 0.136 | 0.409 |
| Heavy metals | **2.30 (1.28, 4.14)** | **0.005** | **0.038** | 2.13 (1.01, 4.50) | 0.048 | 0.166 | 4.63 (1.32, 16.24) | 0.017 | 0.149 | 1.56 (0.85, 2.87) | 0.149 | 0.424 | **3.50 (1.45, 8.47)** | **0.005** | **0.074** |
| Alcohols | 1.23 (0.88, 1.72) | 0.220 | 0.440 | 1.28 (0.69, 2.41) | 0.434 | 0.652 | 0.29 (0.04, 2.17) | 0.227 | 0.533 | 1.28 (0.91, 1.81) | 0.160 | 0.433 | 1.57 (0.82, 2.98) | 0.172 | 0.442 |
| Pesticides/Fumigants | **2.62 (1.42, 4.85)** | **0.002** | **0.025** | 1.23 (0.64, 2.35) | 0.540 | 0.721 | 1.11 (0.38, 3.22) | 0.849 | 0.917 | 1.63 (0.91, 2.92) | 0.099 | 0.372 | 1.85 (0.6, 5.69) | 0.283 | 0.588 |
| Compounds in plastic production | 2.00 (0.98, 4.07) | 0.055 | 0.166 | 1.81 (0.83, 3.93) | 0.136 | 0.350 | 1.38 (0.30, 6.40) | 0.678 | 0.811 | 1.67 (0.86, 3.25) | 0.129 | 0.409 | 2.3 (0.85, 6.17) | 0.100 | 0.372 |
| Dust | **2.18 (1.46, 3.26)** | **<0.001** | **0.004** | 1.12 (0.39, 3.17) | 0.836 | 0.942 | 2.10 (0.9, 4.87) | 0.085 | 0.372 | **2.43 (1.58, 3.75)** | **<0.001** | **0.003** | 1.21 (0.42, 3.51) | 0.722 | 0.811 |
| Emission from fuels | **1.97 (1.23, 3.16)** | **0.005** | **0.038** | 1.31 (0.50, 3.44) | 0.581 | 0.748 | 1.25 (0.48, 3.27) | 0.654 | 0.811 | **2.04 (1.2, 3.49)** | **0.009** | **0.096** | 1.23 (0.47, 3.24) | 0.676 | 0.811 |
| Occupation Carcinogens | 1.04 (0.64, 1.67) | 0.876 | 0.954 | 1.44 (0.82, 2.51) | 0.204 | 0.431 | 2.13 (1.05, 4.32) | 0.036 | 0.218 | 1.12 (0.7, 1.77) | 0.638 | 0.811 | 1.23 (0.57, 2.63) | 0.594 | 0.811 |
| Acids [b] | 1.03 (0.59, 1.79) | 0.927 | 0.954 | 0.96 (0.47, 1.94) | 0.904 | 0.954 | - | - | - | 0.91 (0.54, 1.54) | 0.731 | 0.811 | 1.36 (0.52, 3.56) | 0.530 | 0.811 |
| Alkalis [b] | 1.36 (0.72, 2.56) | 0.337 | 0.606 | 0.74 (0.32, 1.73) | 0.492 | 0.681 | - | - | - | 1.13 (0.64, 1.98) | 0.670 | 0.811 | 1.09 (0.32, 3.68) | 0.888 | 0.940 |
| Stains and varnishes | **2.29 (1.24, 4.25)** | **0.008** | **0.050** | 1.18 (0.58, 2.44) | 0.646 | 0.802 | 2.7 (1.14, 6.43) | 0.025 | 0.190 | 1.33 (0.68, 2.58) | 0.406 | 0.772 | 1.35 (0.39, 4.62) | 0.633 | 0.811 |
| Paints and paint thinners | 1.65 (0.99, 2.75) | 0.055 | 0.166 | 1.37 (0.79, 2.39) | 0.265 | 0.502 | 0.69 (0.24, 2.01) | 0.495 | 0.811 | 1.11 (0.65, 1.89) | 0.704 | 0.811 | **3.25 (1.57, 6.72)** | **0.001** | **0.040** |
| Anesthetics | 1.41 (0.62, 3.17) | 0.412 | 0.645 | 2.11 (0.99, 4.47) | 0.052 | 0.166 | 4.88 (0.47, 50.44) | 0.184 | 0.451 | 1.68 (0.86, 3.26) | 0.127 | 0.409 | 2.82 (0.99, 8.01) | 0.052 | 0.278 |
| Glues and adhesives | 1.70 (1.02, 2.82) | 0.041 | 0.166 | 1.00 (0.55, 1.83) | 0.997 | 0.997 | 1.38 (0.47, 4.11) | 0.559 | 0.811 | 1.49 (0.92, 2.4) | 0.103 | 0.372 | 1.32 (0.54, 3.25) | 0.548 | 0.811 |
| Soldering materials | 0.89 (0.34, 2.33) | 0.816 | 0.942 | 1.56 (0.58, 4.21) | 0.375 | 0.639 | 1.31 (0.27, 6.45) | 0.736 | 0.811 | 1.31 (0.57, 2.97) | 0.524 | 0.811 | 0.43 (0.05, 3.31) | 0.415 | 0.772 |
| Dyes and inks | 1.38 (1.02, 1.85) | 0.034 | 0.166 | 0.95 (0.55, 1.61) | 0.837 | 0.942 | 1.22 (0.57, 2.63) | 0.608 | 0.811 | 1.39 (1.03, 1.89) | 0.033 | 0.218 | 1.23 (0.65, 2.34) | 0.529 | 0.811 |

[a] Reference: never exposure group.

[b] The OR of exposure to acids and alkalis under 1 year were not estimated due to no observations in the case group.

The regression model was adjusted for age, sex, race, highest education, annual household income, and BMI

**Table S3** Association of exposure to specific chemical agents in the workplace with asthma in adults (n=3148)

See Supporting Table S3 that in the .xlsx format

**Table S4** Association between exposure to main chemical groups and multimorbidity in adults (n=3148)

| Chemical groups | Asthma and AR | | | Asthma and COPD | | |
|---|---|---|---|---|---|---|
| | OR (95%CI) | *P* | FDR | OR (95%CI) | *P* | FDR |
| Solvents and degreasers | 1.39 (0.97, 1.98) | 0.069 | 0.144 | 1.26 (0.54, 2.93) | 0.586 | 0.725 |
| Lubricating oils | **1.80 (1.16, 2.80)** | **0.008** | **0.044** | **3.38 (1.61, 7.07)** | **0.001** | **0.010** |
| Cleaning liquids | **1.53 (1.16, 2.03)** | **0.003** | **0.020** | **2.08 (1.12, 3.83)** | **0.019** | **0.066** |
| Heavy metals | **2.36 (1.47, 3.78)** | **<0.001** | **0.005** | **2.9 (1.15, 7.31)** | **0.024** | **0.075** |
| Alcohols | 1.34 (0.97, 1.84) | 0.077 | 0.148 | 1.05 (0.45, 2.45) | 0.907 | 0.976 |
| Pesticides/Fumigants | **1.81 (1.15, 2.87)** | **0.011** | **0.044** | **3.43 (1.57, 7.49)** | **0.002** | **0.012** |
| Compounds in plastic production | **1.90 (1.11, 3.27)** | **0.020** | **0.057** | 2.92 (0.98, 8.71) | 0.054 | 0.126 |
| Dust | **2.04 (1.38, 3.02)** | **<0.001** | **0.005** | **4.05 (2.01, 8.14)** | **<0.001** | **0.001** |
| Emission from fuels | **1.92 (1.25, 2.95)** | **0.003** | **0.020** | **4.47 (2.12, 9.43)** | **<0.001** | **0.001** |
| Occupation Carcinogens | 1.26 (0.87, 1.83) | 0.227 | 0.361 | 2.12 (1.01, 4.45) | 0.048 | 0.118 |
| Acids | 1.07 (0.67, 1.7) | 0.790 | 0.854 | 0.67 (0.16, 2.86) | 0.591 | 0.725 |
| Alkalis | 1.05 (0.61, 1.81) | 0.852 | 0.902 | 0.47 (0.06, 3.53) | 0.466 | 0.663 |
| Stains and varnishes | **1.73 (1.05, 2.84)** | **0.030** | **0.075** | 1.91 (0.77, 4.77) | 0.163 | 0.315 |
| Paints and paint Thinners | 1.50 (1.00, 2.24) | 0.048 | 0.109 | **2.92 (1.44, 5.92)** | **0.003** | **0.017** |
| Anesthetics | **1.97 (1.13, 3.45)** | **0.018** | **0.053** | 2.09 (0.48, 9.06) | 0.326 | 0.550 |
| Glues and adhesives | 1.41 (0.94, 2.14) | 0.100 | 0.186 | **2.26 (1.05, 4.86)** | **0.038** | **0.099** |
| Soldering materials | 1.2 (0.58, 2.48) | 0.621 | 0.729 | 2.31 (0.75, 7.07) | 0.144 | 0.288 |
| Dyes and inks | **1.45 (1.09, 1.92)** | **0.010** | **0.044** | 1.64 (0.87, 3.09) | 0.127 | 0.264 |

Abbreviations: AR, allergic rhinitis; COPD, chronic obstructive pulmonary disease.
The logistic regression model was adjusted for age, sex, race, highest education, annual household income, and BMI.

**Table S5** Association of exposure to specific chemical agents in the workplace with multimorbidity (asthma and AR) in adults (n=3148)

See Supporting Table S5 that in the .xlsx format

**Table S6** Association of exposure to individual chemicals in the workplace with multimorbidity (asthma and COPD) in adults (n=3148)

See Supporting Table S6 that in the .xlsx format

**Table S7** Estimated odd ratios (ORs) for asthma and respiratory multimorbidity in association with polyexposure risk scores (PXS) of known asthmagens and suspected or novel risk agents and the effect modification of PPE in adults (**numeric data for Figure 3**)

| PXS (per 0.1 increase) [a] | N | Asthma (n=307) | | Multimorbidity | | | |
|---|---|---|---|---|---|---|---|
| | | | | Asthma and AR (n=255) | | Asthma and COPD (n=47) | |
| | | OR (95% CI) | *P* int | OR (95% CI) | *P* int | OR (95% CI) | *P* int |
| **Total population** | | | | | | | |
| PXS $_{Known}$ | 3148 | 1.19 (1.11, 1.28) | / | 1.21 (1.12, 1.30) | / | 1.30 (1.13, 1.49) | / |
| PXS $_{Suspected/novel}$ | 3148 | 1.44 (1.27, 1.63) | / | 1.49 (1.31, 1.69) | / | 2.03 (1.64, 2.49) | / |
| **Interaction with PPE** | | | | | | | |
| PXS $_{Known}$ | | | | | | | |
| Wearing gloves | 2518 | | 0.965 | | 0.360 | | 0.553 |
| No | 1205 | 1.21 (1.04, 1.39) | | 1.15 (0.97, 1.34) | | 1.42 (1.10, 1.79) | |
| Yes | 1313 | 1.21 (1.10, 1.32) | | 1.25 (1.14, 1.38) | | 1.29 (1.04,1.60) | |
| Wearing face mask or respirator | 2585 | | 0.559 | | 0.975 | | 0.752 |
| No | 1961 | 1.23 (1.10, 1.36) | | 1.21 (1.08, 1.35) | | 1.30 (1.03, 1.62) | |
| Yes | 624 | 1.17 (1.05, 1.31) | | 1.21 (1.07, 1.35) | | 1.24 (1.00, 1.54) | |
| Wearing protective clothing | 2622 | | 0.257 | | 0.561 | | 0.187 |
| No | 1948 | 1.25 (1.14, 1.38) | | 1.24 (1.11, 1.37) | | 1.33 (1.11, 1.58) | |
| Yes | 674 | 1.15 (1.02, 1.30) | | 1.18 (1.04, 1.34) | | 1.71 (1.23, 2.36) | |
| PXS $_{Suspected\ or\ novel}$ | | | | | | | |
| Wearing gloves | 2518 | | **0.002** | | **0.012** | | **0.028** |
| No | 1205 | 2.01 (1.57, 2.57) | | 1.95 (1.51, 2.51) | | 3.04 (2.05, 4.49) | |
| Yes | 1313 | 1.27 (1.09, 1.49) | | 1.34 (1.14, 1.57) | | 1.77 (1.31, 2.40) | |
| Wearing face mask or respirator | 2585 | | 0.055 | | 0.086 | | 0.992 |
| No | 1961 | 1.65 (1.35, 2.01) | | 1.67 (1.35, 2.05) | | 1.95 (1.30, 2.84) | |
| Yes | 624 | 1.27 (1.07, 1.52) | | 1.31 (1.10, 1.58) | | 1.95 (1.46, 2.62) | |
| Wearing protective clothing | 2622 | | **0.011** | | **0.024** | | 0.629 |
| No | 1948 | 1.73 (1.46, 2.06) | | 1.74 (1.46, 2.08) | | 2.21 (1.67, 2.93) | |
| Yes | 674 | 1.24 (1.02, 1.51) | | 1.28 (1.04, 1.57) | | 2.53 (1.58, 4.06) | |

Abbreviations: AR, allergic rhinitis; COPD, chronic obstructive pulmonary disease; PPE, personal protective equipment.

[a] Weighted standardized risk score (ranging from 0 to 1) was calculated based on the $\beta$ coefficients of chemical exposures in the LASSO regression..

The logistic model was adjusted for age, sex, race, highest education, annual household income, and BMI.

**Table S8** ExWAS results for potential risk chemical agents and their multi-exposure model for adult asthma in the workplace with imputed covariates data (n=3148)

| Predefined Classifications | | Chemical agents | Group | ExWAS results | | LASSO coefficients (β) | |
|---|---|---|---|---|---|---|---|
| | | | | OR (95% CI) | p value/ FDR | Model 1 | Model 2 |
| Known asthmagen [a] | Sensitizer and irritant | Beach | Cleaning liquids | 1.58 (1.22, 2.04) | 0.001 / 0.02 | 0.219 | 0.219 |
| | Sensitizer and irritant | Ammonia | Cleaning liquids | 2.00 (1.40, 2.86) | <0.001 / 0.01 | 0.358 | 0.358 |
| | Sensitizer | Resin | Stains and varnishes | 2.54 (1.21, 5.35) | 0.014 / 0.16 | 0.403 | 0.403 |
| | Sensitizer | Nickel | Heavy metals | 2.65 (1.11, 6.36) | 0.028 / 0.20 | 0.226 | 0.226 |
| Suspected [b] / Novel risk agent | Suspected | Talc | Dust | 3.03 (1.75, 5.24) | <0.001 / 0.01 | 0.778 | 0.820 |
| | Suspected | Carbon monoxide | Emission from fuels | 2.16 (1.27, 3.68) | 0.005 / 0.08 | 0.406 | 0.465 |
| | Suspected | Rubber cement | Glues and adhesives | 1.99 (1.21, 3.26) | 0.007 / 0.11 | 0.249 | 0.323 |
| | Suspected | Vinyl chloride | Plastic production | 3.14 (1.23, 8.05) | 0.017 / 0.16 | 0.400 | 0.476 |
| | Suspected | Acrylic | Stains and varnishes | 2.11 (1.21, 3.70) | 0.009 / 0.12 | 0.156 | - |
| | Novel | Silica | Dust | 3.48 (1.59, 7.65) | 0.002 / 0.04 | 0.504 | 0.589 |
| | Novel | Carbon dioxide | Emission from fuels | 1.93 (1.11, 3.34) | 0.020 / 0.17 | 0.000 | - |
| | Novel | Neoprene | Glues and adhesives | 2.98 (1.23, 7.19) | 0.015 / 0.16 | 0.000 | - |
| | Novel | Nitrous | Anesthetics | 2.29 (1.12, 4.68) | 0.023 / 0.18 | 0.359 | 0.427 |

Abbreviations: ExWAS, exposome-wide association study; OR, odds ratio; FDR, false discovery rate; LASSO, least absolute shrinkage and selection operator.

[a] Known asthmagens were established by Association of Occupational and Environmental Clinics (AOEC) Exposure Code List or World Allergy Organization (WAO); sensitizer or irritant was further determined according to AOEC Exposure Code List and the references of WAO.

[b] Suspected agents were determined by any evidence from previous studies.

All logistic-based ExWAS and LASSO models were adjusted for age, sex, race, highest education, annual household income, and BMI.

To estimate chemical weights of $PXS_{known}$, all known asthmagens selected by ExWAS (FDR<0.20) were included in the model 1 of the LASSO regression, all of which remained in model 2.

To estimate chemical weights of $PXS_{suspected\ or\ novel}$, all suspected or potential novel risk chemical agents selected by ExWAS (FDR<0.20) were included in the model 1 of the LASSO regression, while model 2 further excluded those with minimal contribution (i,e. acrylic, carbon dioxide, and neoprene) in model 1.

**Table S9** Additionally adjusted multi-exposure models for chemical agents and adult asthma in the workplace (n=3148)

| Chemical agents | LASSO coefficients (β) | | |
|---|---|---|---|
| | Main model (chemical with FDR<0.20) | Main model + other chemicals (*P*<0.05) | Main model + other chemicals (*FDR*<0.30) |
| Beach | 0.192 | - | 0.215 |
| Ammonia | 0.413 | - | 0.423 |
| Resin | 0.505 | - | 0.572 |
| Nickel | 0.221 | - | 0.303 |
| Talc | 0.858 | 0.852 | 0.766 |
| Carbon monoxide | 0.502 | 0.540 | 0.414 |
| Rubber cement | 0.272 | 0.259 | 0.163 |
| Vinyl chloride | 0.481 | 0.504 | 0.400 |
| Silica | 0.585 | 0.641 | 0.499 |
| Nitrous | 0.460 | 0.427 | 0.334 |

Abbreviations: FDR, false discovery rate; LASSO, least absolute shrinkage and selection operator.

The main LASSO model: chemical with FDR<0.20 in the main analysis and covariates, including age, sex, race, highest education, annual household income, and BMI.

Additional model 1: the main LASSO model + chemical agents with P<0.05 in the ExWAS (no additional agents were identified for known asthmagens; toner, mercury, and acetone were adjusted in the model of suspected or novel risk agents).

Additional model 2: the main LASSO model + chemical agents with FDR<0.30 in the ExWAS (ozone and styrene were adjusted in the model of known asthmagens; toner, india ink, mercury, acetone, magnesium-oh, rock dust, inkjet, enamel, shellac, and white glue were adjusted in the model of suspected or novel risk agents).

**Table S10** Estimated odd ratios (ORs) for asthma and respiratory multimorbidity in association with high risk (top 10% PXS) of known asthmagens and suspected suspected/novel risk agents and the effect modification of PPE in adults

| PXS (top 10%) | N | Asthma (n=307) | | Multimorbidity | | | |
|---|---|---|---|---|---|---|---|
| | | | | Asthma and AR (n=255) | | Asthma and COPD (n=47) | |
| | | OR (95% CI) | *P* int | OR (95% CI) | *P* int | OR (95% CI) | *P* int |
| **Total population** | | | | | | | |
| PXS $_{\text{Known}}$ | 3148 | 1.96 (1.37, 2.75) | / | 2.02 (1.38, 2.91) | / | 2.56 (1.19, 5.09) | / |
| PXS $_{\text{Suspected or novel}}$ | 3148 | 2.61 (1.82, 3.69) | / | 2.81 (1.92, 4.04) | / | 7.75 (3.88, 15.21) | / |
| **Interaction with PPE** | | | | | | | |
| PXS $_{\text{Known}}$ | | | | | | | |
| Wearing gloves | 2518 | | 0.706 | | 0.295 | | 0.735 |
| No | 1205 | 1.76 (0.86, 3.35) | | 1.48 (0.66, 2.99) | | 2.84 (0.77, 8.38) | |
| Yes | 1313 | 2.06 (1.31, 3.26) | | 2.37 (1.47, 3.84) | | 2.11 (0.60, 7.39) | |
| Wearing face mask or respirator | 2585 | | 0.62 | | 0.944 | | 0.960 |
| No | 1961 | 2.13 (1.31, 3.36) | | 2.04 (1.20, 3.34) | | 2.38 (0.73, 6.57) | |
| Yes | 624 | 1.76 (0.97, 3.19) | | 1.98 (1.07, 3.68) | | 2.29 (0.66, 7.96) | |
| Wearing protective clothing | 2622 | | 0.322 | | 0.630 | | 0.713 |
| No | 1948 | 2.35 (1.49, 3.62) | | 2.2 (1.34, 3.52) | | 3.34 (1.39, 7.45) | |
| Yes | 674 | 1.59 (0.84, 3.00) | | 1.8 (0.93 ,3.5) | | 5.01 (0.69, 36.45) | |
| PXS $_{\text{Suspected or novel}}$ | | | | | | | |
| Wearing gloves | 2518 | | **0.002** | | **0.016** | | 0.186 |
| No | 1205 | 5.46 (3.03, 9.63) | | 5.23 (2.81, 9.47) | | 14.41 (5.18, 39.64) | |
| Yes | 1313 | 1.70 (1.03, 2.81) | | 2.00 (1.20, 3.36) | | 5.11 (1.50, 17.43) | |
| Wearing face mask or respirator | 2585 | | 0.058 | | 0.061 | | 0.803 |
| No | 1961 | 3.40 (2.09, 5.41) | | 3.65 (2.18, 5.95) | | 8.16 (2.84, 21.92) | |
| Yes | 624 | 1.62 (0.88, 2.98) | | 1.71 (0.91,3.2) | | 6.72 (1.98, 22.75) | |
| Wearing protective clothing | 2622 | | 0.056 | | 0.097 | | 0.676 |
| No | 1948 | 3.79 (2.38, 5.91) | | 3.88 (2.37, 6.19) | | 10.29 (4.56, 22.79) | |
| Yes | 674 | 1.79 (0.96, 3.36) | | 1.97 (1.03, 3.78) | | 17.23 (1.72, 172.47) | |

Abbreviations: PXS, polyexposure risk score; AR, allergic rhinitis; COPD, chronic obstructive pulmonary disease; PPE, personal protective equipment.
The logistic model was adjusted for age, sex, race, highest education, annual household income, and BMI.

**Table S11** Association of polyexposure risk scores (PXS) of chemical exposures and related protective measures with asthma symptom control and respiratory multimorbidity in patients with asthma (N=307)

| | Asthma symptom control in the last 14 days [β (95%CI)] | | | Uncontrolled asthma symptoms [c] [OR (95%CI)] |
|---|---|---|---|---|
| | Night waking days | Activities limitation days | Wheezing days | |
| **Mean (SD) / n (%)** | 0.68 (2.03) | 1.34 (2.94) | 2.18 (3.65) | 126 (41.0%) |
| **PXS $_{Known}$** | | | | |
| Continuous (per 0.1 increase) | **0.16(0.04, 0.28)** ** | **0.17(0.00, 0.34)** * | 0.14(-0.07, 0.36) | 1.09(0.96, 1.25) |
| High risk (top 10% score) | 0.47(-0.16, 1.11) | 0.27(-0.61, 1.16) | -0.04(-1.18, 1.10) | 0.96(0.48, 1.90) |
| **PXS $_{Suspected / novel}$** | | | | |
| Continuous (per 0.1 increase) | **0.31(0.10, 0.52)** ** | **0.33(0.04, 0.62)** * | **0.43(0.05, 0.80)** * | **1.41(1.10, 1.85)** ** |
| High risk (top 10% score) | **0.96(0.32, 1.60)** ** | **1.14(0.26, 2.03)** * | **1.76(0.63, 2.89)** ** | **2.62(1.31, 5.31)** ** |

[a] Weighted standardized risk score (ranging from 0 to 1) was calculated based on the $\beta$ coefficients from the Lasso regression. The regression model was adjusted for age, sex, race, highest education, annual household income, and BMI.
[b] Adjusted for age, sex, race, highest education, annual household income, BMI.
[c] Uncontrolled asthma symptoms: experiencing night waking and activity limitation due to asthma once a week and having asthma symptoms two days a week.
*p<0.05, ** p<0.01, ***p<0.001

**Table S12** The modification effect of sex on the association of polyexposure risk scores (PXS) with asthma and respiratory multimorbidity in adults

| PXS | Current asthma (n=307) | | Multimorbidity | | | |
|---|---|---|---|---|---|---|
| | | | Asthma and AR (n=255) | | Asthma and COPD (n=47) | |
| | OR (95% CI) | *P* int | OR (95% CI) | *P* int | OR (95% CI) | *P* int |
| **Continuous (per 0.1 increase)** | | | | | | |
| PXS $_{Known}$ | | 0.716 | | 0.960 | | 0.271 |
| Male (n=945) | 1.17 (1.00, 1.33) | | 1.20 (1.03, 1.39) | | 1.09 (0.71, 1.49) | |
| Female (n=2179) | 1.20 (1.11, 1.31) | | 1.21 (1.11, 1.32) | | 1.36 (1.17, 1.58) | |
| PXS $_{Suspected\ or\ novel}$ | | 0.078 | | **0.047** | | 0.097 |
| Male (n=945) | 1.27 (1.02, 1.53) | | 1.26 (1.00, 1.55) | | 1.61 (1.04, 2.25) | |
| Female (n=2179) | 1.60 (1.35, 1.88) | | 1.67 (1.41, 1.97) | | 2.36 (1.80, 3.10) | |
| **High risk (top 10% score)** | | | | | | |
| PXS $_{Known}$ | | 0.810 | | 0.863 | | 0.501 |
| Male (n=945) | 1.81 (0.83, 3.62) | | 2.15 (0.93, 4.51) | | 1.60 (0.24, 6.66) | |
| Female (n=2179) | 2.00 (1.35, 2.98) | | 1.99 (1.31, 3.03) | | 2.95 (1.31, 6.61) | |
| PXS $_{Suspected\ or\ novel}$ | | 0.403 | | 0.257 | | 0.234 |
| Male (n=945) | 2.05 (1.01, 3.92) | | 1.94 (0.87, 3.99) | | 3.93 (0.97, 14.45) | |
| Female (n=2179) | 2.87 (1.89, 4.35) | | 3.20 (2.09, 4.90) | | 9.81 (4.56, 21.12) | |

Abbreviations: AR, allergic rhinitis; COPD, chronic obstructive pulmonary disease.
The logistic model was adjusted for age, sex, race, highest education, annual household income, and BMI.

**Table S13** Workplace characteristics and asthma and respiratory multimorbidity in adults

| Workplace characteristics | n / N | OR (95%CI) | | |
|---|---|---|---|---|
| | | Asthma | Asthma and AR | Asthma and COPD |
| Workspace repairs | 657/3047 | 1.16(0.86, 1.55) | 1.25(0.91, 1.70) | 1.15(0.50, 2.40) |
| Floor materials | | | | |
| Concrete | 1045/3148 | 0.88(0.67, 1.15) | 0.89(0.66, 1.18) | 0.94(0.48, 1.75) |
| Wood | 370/3148 | 1.06(0.71, 1.53) | 1.09(0.72, 1.62) | 1.74(0.76, 3.59) |
| Vinyl | 823/3148 | 1.09(0.82, 1.43) | 1.10(0.81, 1.47) | 0.94(0.43, 1.88) |
| Carpet | 1552/3148 | 1.10(0.85, 1.41) | 1.11(0.84, 1.45) | 1.02(0.54, 1.90) |
| Wall materials | | | | |
| Textile | 461/3028 | 1.14(0.81, 1.58) | 1.31(0.91, 1.83) | 0.65(0.22, 1.55) |
| Plastic | 301/3148 | 1.05(0.68, 1.55) | 1.08(0.69, 1.64) | 0.64(0.15, 1.82) |

Abbreviations: PXS, polyexposure risk score; AR, allergic rhinitis; COPD, chronic obstructive pulmonary disease.

The regression model was adjusted for age, sex, race, highest education, annual household income, and BMI.

**Table S14** Demographic characteristics, asthma, and work environment exposures and in the U.S. adults, NHANES 2007–2012 (n = 14174)

| Weighted mean / % (SE) [b] | All | Years 2007-2008 | Years 2009-2010 | Years 2011-2012 |
|---|---|---|---|---|
| | (N=14174) [a] | (n=4770) [a] | (n=4946) [a] | (n=4458) [a] |
| **Demographic characteristics** | | | | |
| Age, year | 44.84 (0.37) | 44.28 (0.53) | 45.03 (0.36) | 45.19 (0.87) |
| Sex | | | | |
| Male | 49.48 (0.46) | 49.66 (0.73) | 49.21 (0.68) | 49.56 (0.94) |
| Female | 50.52 (0.46) | 50.34 (0.73) | 50.79 (0.68) | 50.44 (0.94) |
| Race/ethnicity | | | | |
| Mexican American | 7.88 (1.02) | 69.96 (3.62) | 69.78 (3.14) | 67.63 (3.80) |
| Non-Hispanic White | 11.82 (0.92) | 8.09 (1.35) | 8.08 (2.14) | 7.50 (1.74) |
| Non-Hispanic Black | 69.10 (2.04) | 10.93 (1.75) | 10.93 (1.56) | 13.53 (1.44) |
| Others | 11.20 (1.05) | 11.03 (2.04) | 11.21 (0.83) | 11.34 (2.25) |
| Highest education | | | | |
| Below high school | 14.37 (0.69) | 13.53 (1.07) | 12.95 (0.90) | 16.54 (1.52) |
| High school or equivalent | 26.06 (0.86) | 27.24 (1.64) | 26.03 (0.85) | 24.95 (1.82) |
| Some College or AA degree | 35.32 (1.19) | 37.22 (1.84) | 36.79 (1.84) | 32.07 (2.32) |
| College graduate or above | 24.26 (1.44) | 22.02 (2.03) | 24.23 (1.38) | 26.44 (3.44) |
| Annual household income | | | | |
| Less than $20,000 | 18.82 (0.79) | 21.26 (1.38) | 18.88 (0.96) | 16.44 (1.59) |
| $20,000 to 44,999 | 21.83 (0.77) | 24.18 (1.36) | 22.56 (1.16) | 18.88 (1.43) |
| $45,000 to 99,999 | 30.28 (0.64) | 28.67 (0.93) | 29.98 (0.85) | 32.11 (1.46) |
| $100,000 or above | 29.07 (1.24) | 25.90 (2.03) | 28.59 (1.45) | 32.57 (2.61) |
| BMI, $kg/m^2$ | 28.70 (0.11) | 28.51 (0.17) | 28.84 (0.14) | 28.75 (0.23) |
| **Outcome and exposures** | | | | |
| Current asthma | | | | |
| No | 92.13 (0.48) | 92.45 (0.81) | 92.46 (0.55) | 91.49 (1.05) |
| Yes | 7.87 (0.48) | 7.55 (0.81) | 7.54 (0.55) | 8.51 (1.05) |
| Mineral dust | | | | |
| No | 69.14 (0.78) | 67.97 (1.57) | 69.44 (1.42) | 69.96 (0.99) |
| Yes | 30.86 (0.78) | 32.03 (1.57) | 30.56 (1.42) | 30.04 (0.99) |
| Exhaust fumes | | | | |
| No | 74.96 (0.74) | 74.04 (1.39) | 75.15 (1.41) | 75.66 (0.97) |
| Yes | 25.04 (0.74) | 25.96 (1.39) | 24.85 (1.41) | 24.34 (0.97) |
| Other fumes | | | | |
| No | 68.38 (0.78) | 67.45 (1.25) | 68.40 (1.06) | 69.24 (1.61) |
| Yes | 31.62 (0.78) | 32.55 (1.25) | 31.60 (1.06) | 30.76 (1.61) |

[a] Unweighted sample size.

[b] Data presented are weighted mean (SE) for age and BMI. For other categorical variables, weighted percentages (SE) are presented.

**Table S15** Association between asthma and work environment exposures and in the U.S. adults, NHANES 2007–2012 (n = 14174)

| | All survey cycles [OR (95%CI)] | Individual cycle [OR (95%CI)] | | |
|---|---|---|---|---|
| | | 2007-2008 | 2009-2010 | 2011-2012 |
| Crude model | | | | |
| Mineral dust | 0.97 (0.83, 1.14) | 0.82 (0.57, 1.16) | 1.09 (0.87, 1.37) | 1.03 (0.76, 1.39) |
| Exhaust fumes | 0.99 (0.81, 1.20) | 0.92 (0.71, 1.20) | 1.07 (0.73, 1.56) | 0.98 (0.65, 1.49) |
| Other fumes | 1.09 (0.94, 1.27) | 1.02 (0.81, 1.28) | 1.13 (0.83, 1.53) | 1.13 (0.86, 1.49) |
| Adjusted model | | | | |
| Mineral dust | **1.23 (1.01, 1.50)** * | 1.01 (0.59, 1.71) | 1.46 (0.93, 2.31) | 1.22 (0.73, 2.02) |
| Exhaust fumes | 1.19 (0.95, 1.48) | 1.10 (0.73, 1.63) | 1.37 (0.72, 2.61) | 1.13 (0.63, 2.05) |
| Other fumes | **1.25 (1.05, 1.48)** * | 1.17 (0.78, 1.77) | 1.29 (0.78, 2.15) | 1.29 (0.85, 1.95) |

Odds ratios (ORs) were estimated in the survey-weighted generalized linear regression with a logit link.
Crude model was not adjusted for covariates; adjusted model included age, sex, race/ethnicity, the highest education level, annual household income, BMI, and the survey cycle.
Mineral dust: ever exposure to dust from rock, sand, concrete, coal, asbestos, silica or soil in any job
Exhaust fumes: ever exposure to exhaust fumes from trucks, buses, heavy machinery or diesel engines
Other fumes: ever exposure to any other gases, vapors or fumes, such as vapors from paints, cleaning products, glues, solvents, and acids; or welding/soldering fumes.
*$p<0.05$

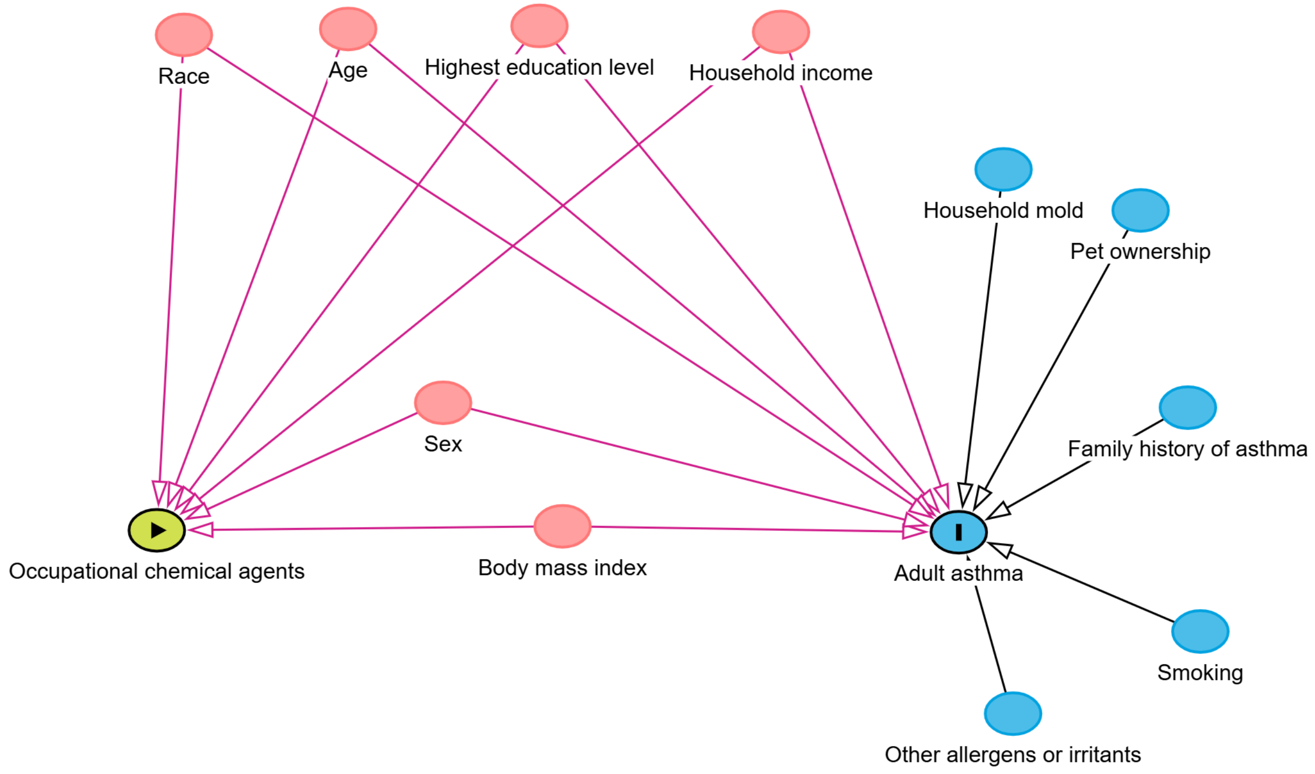


**Figure S1** Directed acyclic graph (DAG) for the hypothesized relationship between occupational chemical exposures and adult asthma in the PEGS participants

Green circle: ancestor of exposure; Blue circle: ancestor of outcome; Red circle: ancestor of exposure and outcome; Green line: causal path; Red line: biasing path.

The minimal sufficient adjustment sets for estimating the total effect of occupational chemical agents on adult asthma:  Age, Body mass index, Highest education level, Household income, Race, Sex

Graph was drawn with using DAGitty version 3.1 (http://dagitty.net/)

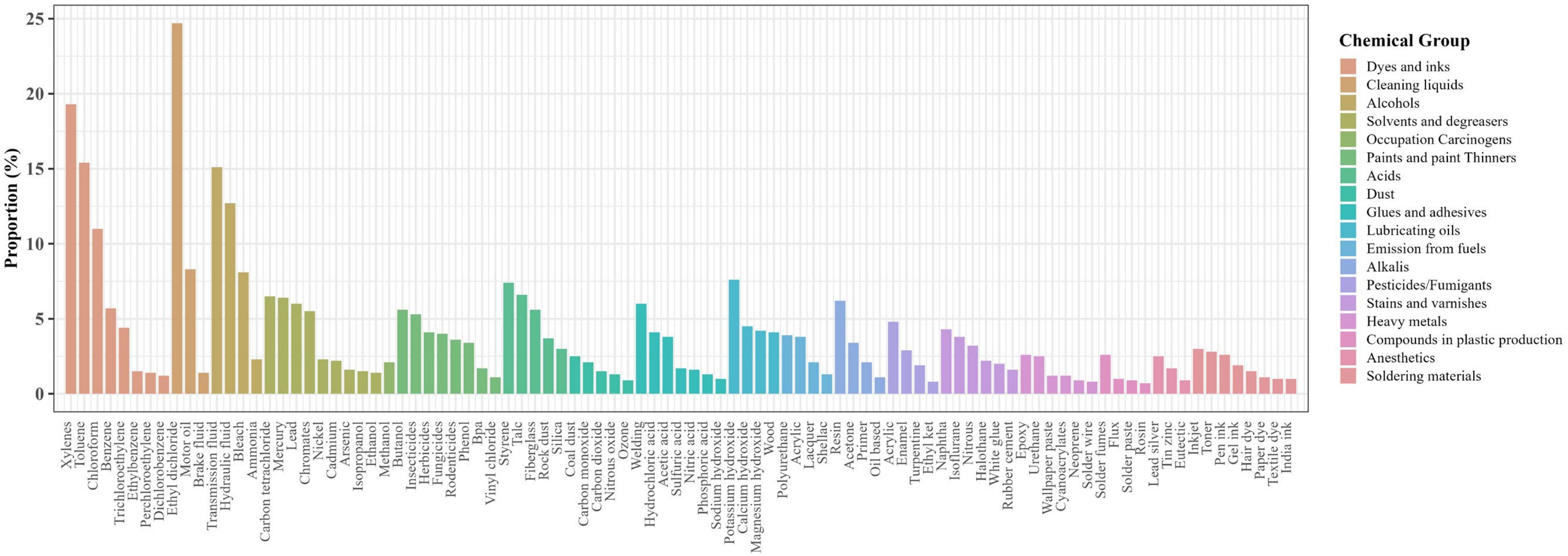


**Figure S2** Proportion of adults ever exposure to individual chemicals within eighteen groups in the workplace (n=3148)

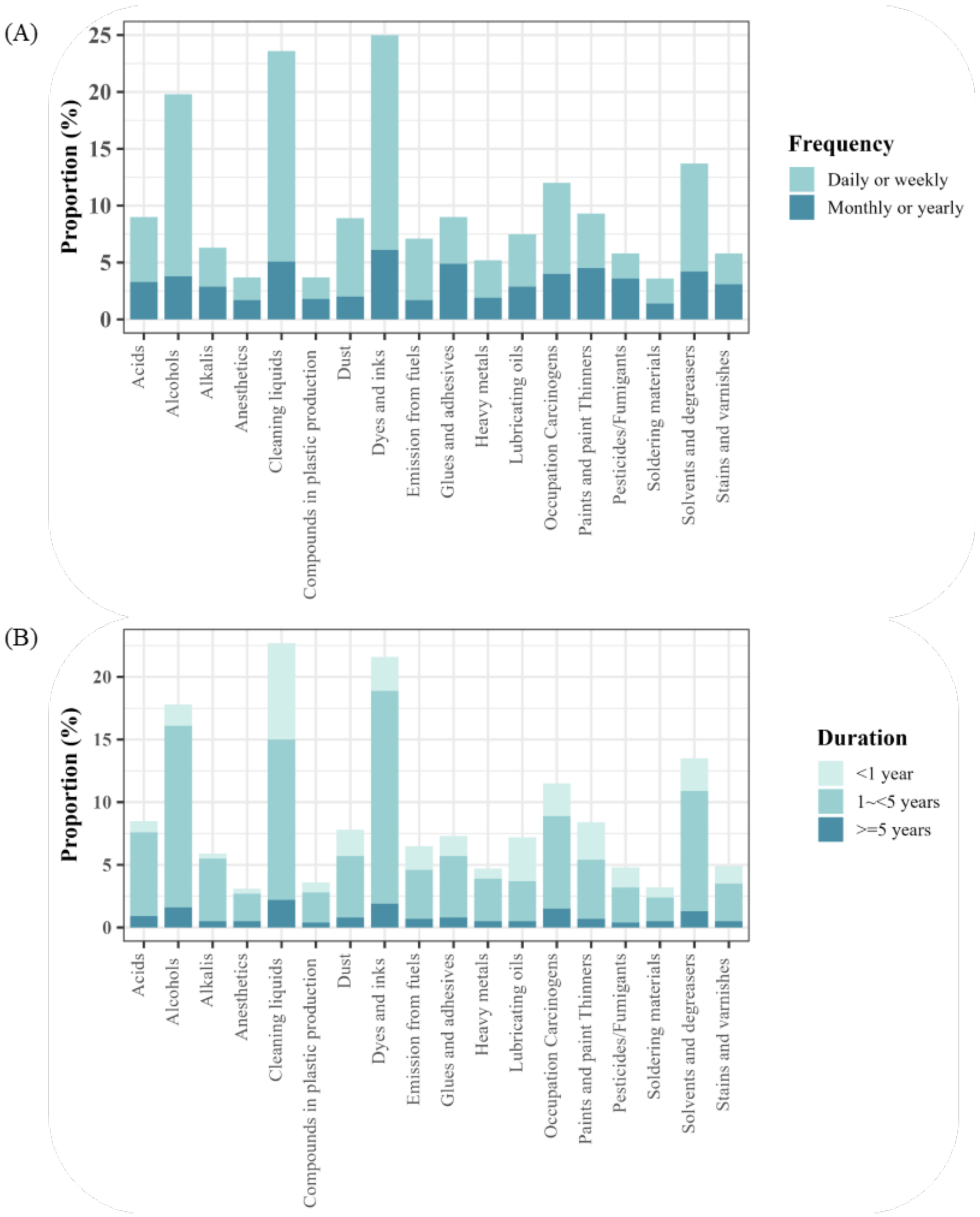


**Figure S3** Exposure frequency (A) and duration (B) to chemical groups at work in adults (n=3148)

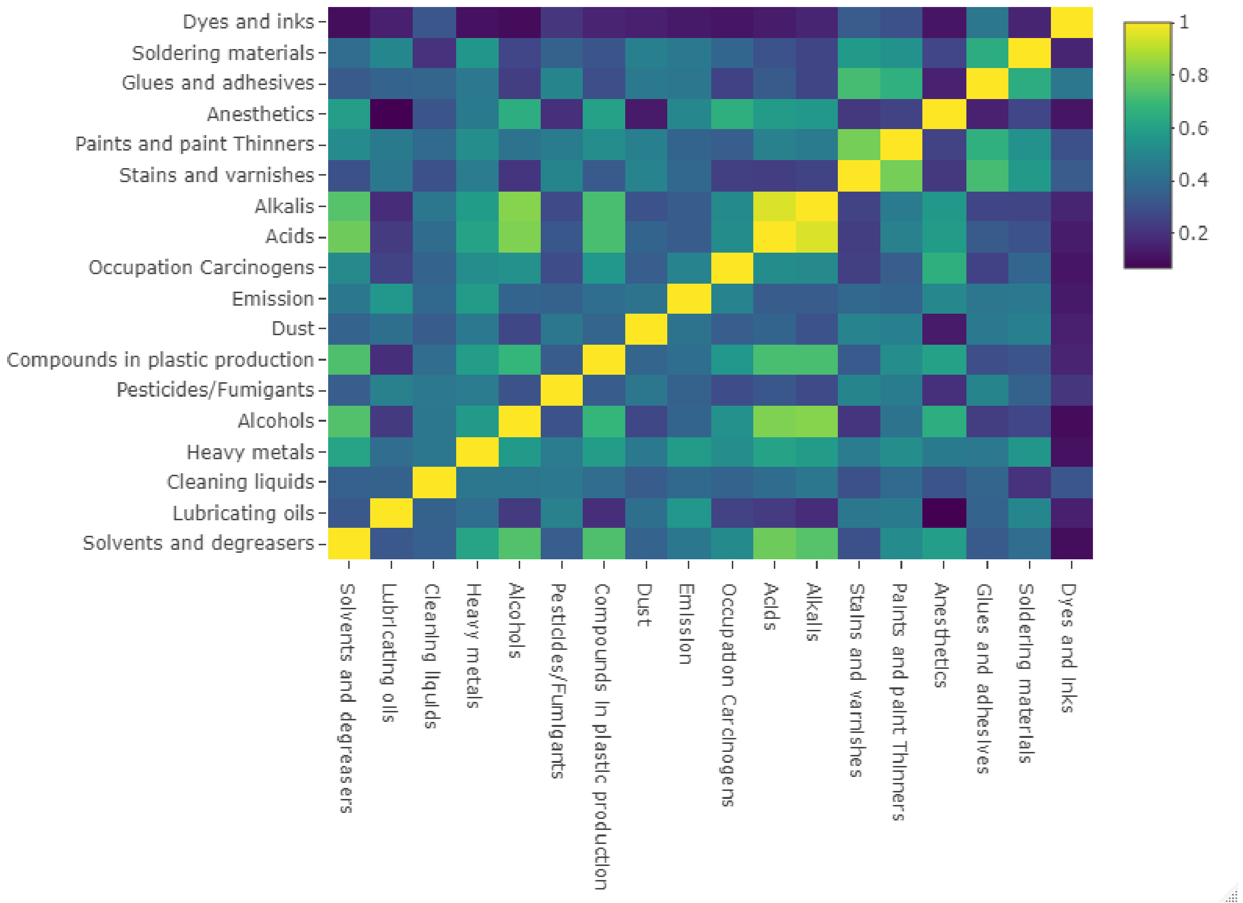


**Figure S4** Pairwise correlation among chemical exposure groups

The interactive plots showing the pairwise correlation between chemical groups and specific chemicals are also displayed in the shiny app: https://lihangyu.shinyapps.io/asthma_shiny
